# Accelerated waning of the humoral response to SARS-CoV-2 vaccines in obesity

**DOI:** 10.1101/2022.06.09.22276196

**Authors:** Agatha A. van der Klaauw, Emily C. Horner, Pehuén Pereyra-Gerber, Utkarsh Agrawal, William S. Foster, Sarah Spencer, Bensi Vergese, Miriam Smith, Elana Henning, Isobel D. Ramsay, Jack A. Smith, Stephane M. Guillaume, Hayley J. Sharpe, Iain M. Hay, Sam Thompson, Silvia Innocentin, Lucy H. Booth, Chris Robertson, Colin McCowan, Thomas E. Mulroney, Martin J. O’Reilly, Thevinya P. Gurugama, Lihinya P. Gurugama, Maria A. Rust, Alex Ferreira, Soraya Ebrahimi, Lourdes Ceron-Gutierrez, Jacopo Scotucci, Barbara Kronsteiner, Susanna J. Dunachie, Paul Klenerman, PITCH Consortium, Adrian J. Park, Francesco Rubino, Hannah Stark, Nathalie Kingston, Rainer Doffinger, Michelle A. Linterman, Nicholas J. Matheson, Aziz Sheikh, I. Sadaf Farooqi, James E. D. Thaventhiran

**Affiliations:** University of Cambridge Metabolic Research Laboratories and NIHR Cambridge Biomedical Research Centre, Wellcome-Medical Research Council (MRC) Institute of Metabolic Science; MRC Toxicology Unit, University of Cambridge, Cambridge, UK; Cambridge Institute of Therapeutic Immunology and Infectious Disease, University of Cambridge, Cambridge, UK; Department of Medicine, University of Cambridge, Cambridge, UK; Cambridge Institute for Medical Research, University of Cambridge, Cambridge, UK; NIHR Cambridge Clinical Research Facility, Cambridge University Hospitals NHS Foundation Trust, Cambridge, UK; Departments of Immunology, Cambridge University Hospitals NHS Foundation Trust, Cambridge, UK; Clinical Biochemistry, Cambridge University Hospitals NHS Foundation Trust, Cambridge, UK; Infectious Diseases, Cambridge University Hospitals NHS Foundation Trust, Cambridge, UK; NIHR BioResource, Cambridge University Hospitals NHS Foundation Trust, Cambridge, UK; NHS Blood and Transplant, Cambridge, UK; Babraham Institute, Babraham Research Campus, Cambridge, UK; Department of Mathematics and Statistics, University of Strathclyde, UK; School of Medicine, University of St. Andrew’s, UK; Usher Institute, University of Edinburgh, UK; Nuffield Department of Clinical Medicine, University of Oxford, UK; Department of Diabetes, King’s College London and Kings College Hospital NHS Foundation Trust, London, UK

**Keywords:** Obesity, SARS-CoV-2, Covid-19, vaccine, humoral immunity

## Abstract

Obesity is associated with an increased risk of severe Covid-19. However, the effectiveness of SARS-CoV-2 vaccines in people with obesity is unknown. Here we studied the relationship between body mass index (BMI), hospitalization and mortality due to Covid-19 amongst 3.5 million people in Scotland. Vaccinated people with severe obesity (BMI>40 kg/m^2^) were significantly more likely to experience hospitalization or death from Covid-19. Excess risk increased with time since vaccination. To investigate the underlying mechanisms, we conducted a prospective longitudinal study of the immune response in a clinical cohort of vaccinated people with severe obesity. Compared with normal weight people, six months after their second vaccine dose, significantly more people with severe obesity had unquantifiable titres of neutralizing antibody against authentic SARS-CoV-2 virus, reduced frequencies of antigen-experienced SARS-CoV-2 Spike-binding B cells, and a dissociation between anti-Spike antibody levels and neutralizing capacity. Neutralizing capacity was restored by a third dose of vaccine, but again declined more rapidly in people with severe obesity. We demonstrate that waning of SARS-CoV-2 vaccine-induced humoral immunity is accelerated in people with severe obesity and associated with increased hospitalization and mortality from breakthrough infections. Given the prevalence of obesity, our findings have significant implications for global public health.

## Main

Globally, obesity (defined as a body mass index (BMI)>30 kg/m^2^) is a major risk factor for severe Covid-19^1^. Severe obesity (BMI>40 kg/m^2^), which affects 3% of the population in the UK and 9% in the US (www.worldobesity.org), is associated with a 90% higher risk of death from Covid-19^2^. Obesity is associated with type 2 diabetes mellitus, hypertension, chronic kidney disease (CKD) and heart failure, co-morbidities which independently increase the risk of severe Covid-19^3^.

SARS-CoV-2 vaccines reduce the risk of symptomatic infection, hospitalization and mortality due to Covid-19^4–8^. They generate antibodies against the Spike (S) protein of SARS-CoV-2, comprising S1 and S2 subunits; S1 contains the receptor binding domain (RBD), which mediates binding of the virus to angiotensin converting enzyme-2 (ACE-2) on host cells. The RBD is the main target for SARS-CoV-2 neutralizing antibodies, which inhibit viral replication *in vitro* and correlate with protection against infection *in vivo*^4–6^. As well as neutralizing antibodies, non-neutralizing antibodies and cellular immunity contribute to protection, particularly against severe Covid-19. Immunity acquired after two doses of vaccine wanes over 6-9 months; many countries have elected to administer booster doses to maintain immune protection, particularly in the elderly and immunocompromised^9,10^.

People with obesity have impaired immune responses to conventional influenza, rabies and hepatitis vaccines^11–14^, however, the impact of obesity on their responses to mRNA and adenoviral vectored vaccines is not known. Some studies have suggested that following SARS-CoV-2 vaccination, antibody titres may be lower in people with obesity^15–20^. One possible explanation is the impact of needle length on vaccine dosing in people with obesity^21^. To date, longitudinal studies to investigate the duration of protection following SARS-CoV-2 vaccination in people with obesity have not been performed.

### Hospitalization and death due to Covid-19 in vaccinated people

To investigate the real-world effectiveness of SARS-CoV-2 vaccination, we used the EAVE II surveillance platform which draws on near real-time nationwide health care data for 5.4 million individuals (∼99%) in Scotland, UK^22–25^. We interrogated data on 3,522,331 individuals aged 18 and over who received a second dose (of the primary vaccination schedule) or a third booster dose of vaccine and were followed up until hospitalization, death or the end of the study. Body mass index (BMI, weight in kg/height in metres squared) was recorded for 1,734,710 (49.2%) individuals. We first examined the impact of BMI on Covid-19 related hospitalization and mortality ≥14 days after receiving a second dose of either Pfizer-BioNTech BNT162b2 mRNA or AstraZeneca ChAdOx1 nCoV-19 vaccines. Between September 14, 2020 and March 19, 2022, there were 10,983 people (0.3%, 6.0 events per 1000 person-years) who had a severe Covid-19 outcome: 9,733 individuals were hospitalized and 2,207 individuals died due to Covid-19 (957 individuals were hospitalized before their death). People with severe obesity (BMI>40 kg/m^2^) were at increased risk of severe Covid-19 outcomes following a second vaccine dose, compared to those with a BMI in the normal range, with an adjusted Rate Ratio (aRR) of 1.76 (95% Confidence Intervals (CI), 1.60-1.94) after adjusting for age, sex and socioeconomic status (Methods). A modest increase in risk was also seen in people who were obese (BMI 30-40 kg/m^2^) and those who were underweight (BMI <18.5 kg/m^2^) (aRR 1.11, 95% CI 1.05-1.18 and aRR 1.28, 95% CI 1.12-1.47, respectively) (Table S1 in the Extended Data). People with obesity and severe obesity were at higher risk of hospitalization or death from Covid-19 after both a second (Fig. 1) and third (booster) dose (Fig. S1 in the Extended Data). Breakthrough infections after the second vaccine dose presented sooner in people with severe obesity (10 weeks) and obesity (15 weeks) than in normal weight people (20 weeks) (Fig. 1). To interrogate whether vaccine effectiveness differed over time in people with severe obesity, an interaction test (comparing one model with the interaction terms in it and then a second model without the interaction terms) was performed. This indicated that vaccine effectiveness over time differed across BMI groups with more rapid loss of protection in those with increased BMI (P<0.001).

**Figure 1.**
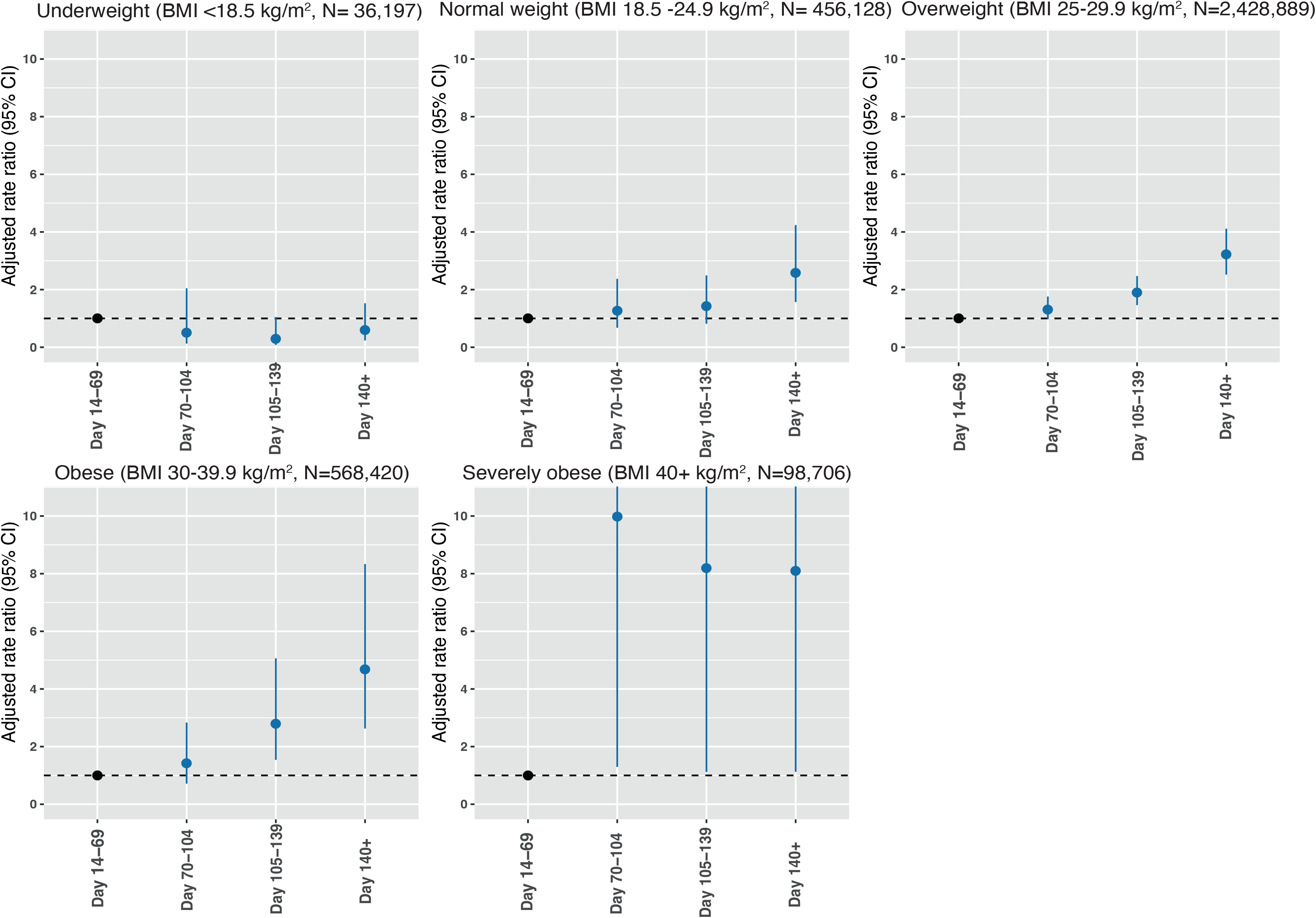
Risk of severe Covid-19 outcomes after primary vaccination and relationship with body mass index. Panels depict the adjusted rate ratios (aRR) for hospitalization or death (severe Covid-19 outcomes) with time after the second SARS-CoV-2 vaccine dose for people in each BMI (body mass index) category in the EAVE-II cohort, Scotland. ARRs are calculated against baseline risk at 14-69 days after the second vaccine dose. Error bars indicate 95% confidence intervals. The number (N) of people in each BMI category is indicated. Breakthrough infections after the second vaccine dose presented more quickly in people with severe obesity (10 weeks) and obesity (15 weeks) than in normal weight people (20 weeks).

Vaccinated individuals who were severely obese and also had type 2 diabetes were at increased risk of admission to hospital or death due to Covid-19 (aRR 1.43, 95% CI 1.17-1.74, Fig. S1f in the Extended Data). A diagnosis of type 2 diabetes was independently associated with an increased risk of a severe Covid-19 outcome despite vaccination (aRR 1.13, 95% CI 1.07-1.19), although this was less than the risk associated with severe obesity. The aRR for type 2 diabetes was reduced slightly after adjusting for BMI (1.06, 95% CI 1.00-1.12). Further studies will be needed to test whether hyperglycaemia modifies the risk associated with severe obesity. Cardiovascular disease (stroke, peripheral vascular disease or coronary heart disease), heart failure, asthma, or chronic kidney disease were not significantly associated with a further increase in risk in vaccinated people (Fig. S1f in the Extended Data).

### Humoral and cellular immunity after primary vaccination

To investigate the mechanisms underlying reduced vaccine efficacy in people with higher BMI, we performed detailed prospective longitudinal immunophenotyping of a clinical cohort of people with severe obesity (n=28) and normal weight controls (n=41) in Cambridge, UK (SCORPIO study) (Fig. 2a and Table S2 in the Extended Data). All participants had received a two-dose primary course of SARS-CoV-2 vaccine approximately 6 months prior to study enrolment (Fig. 2a). As prior natural SARS-CoV-2 infection enhances subsequent vaccination responses, people with detectable anti-nucleocapsid antibodies (n= 2 with severe obesity; n= 1 normal weight control) were excluded. Mean levels of anti-Spike and anti-RBD IgG antibodies were comparable between people with severe obesity and normal weight controls six months after the second vaccine dose (Fig. 2b and Fig. S2a in the Extended Data). In contrast, the function of these antibodies, measured by their ability to neutralize authentic SARS-CoV-2 viral infection (NT50; neutralizing titres at 50% inhibition), was reduced in people with severe obesity (Fig. 2c). In fact, 56% of individuals with severe obesity had unquantifiable neutralizing capacity, compared to 12% of normal weight controls (P=0.0007, Fisher’s exact test, Fig. 2d). The observed dissociation between Spike antibody levels and neutralizing capacity could be a consequence of lower antibody affinity, or differential antibody reactivity to non-neutralizing epitopes of the Spike protein. Here, normal levels of RBD-binding antibodies indicate adequate capacity for antibody production against neutralizing epitopes, suggesting a lower affinity of SARS-CoV-2 antibodies in people with severe obesity. Baseline plasma glucose, leptin levels or the presence type 2 diabetes did not correlate with neutralizing capacity in people with severe obesity (Fig. S2b-e in the Extended Data).

**Figure 2.**
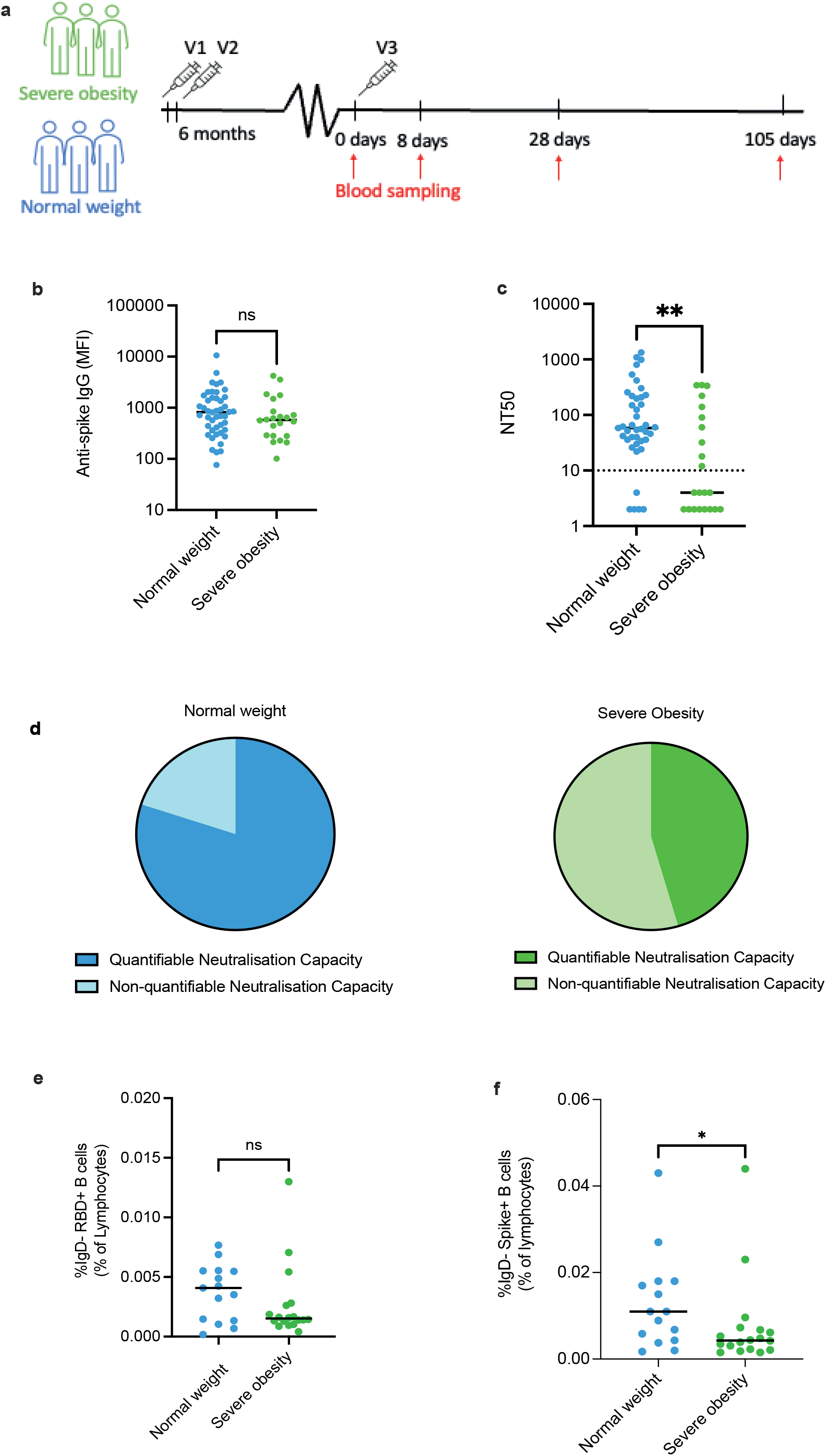
SARS-CoV-2 vaccine-induced immunity in people with severe obesity and normal weight people six months after primary vaccination. **a,** Detailed longitudinal immunophenotyping studies were performed on people with severe obesity (green) and normal weight controls (blue). Samples were obtained six months after the second dose of SARS-CoV-2 vaccine (V2) administered as part of their primary vaccination course and at several time-points after the third dose (V3) as indicated. **b,** shows levels of anti-Spike IgG antibodies (MFI, mean fluorescence intensity). **c**, shows neutralizing titres at 50% inhibition (NT50) against wild type SARS-CoV-2, with the dotted line indicating the limit of quantitation. **d**, shows the proportion of people in both groups with unquantifiable and quantifiable titres of neutralizing antibodies. **e**, shows the frequency of antigen-experienced (IgD-) Receptor Binding Domain binding (RBD+) and Spike-binding (Spike+) B cells, 6 months after the primary vaccination course; data expressed as a percentage (%) of the total number of lymphocytes. Panels **b, c, e, f**; horizontal bars indicate the median; ns, not significant, *P<0.05, **P<0.01.

Suboptimal antibody responses may be enhanced by activating memory B-cells, which can rapidly differentiate into antibody-producing plasma cells after booster immunisation. Consistent with the impaired neutralizing antibody response in people with severe obesity, we found a trend towards a reduced frequency of antigen-experienced (IgD-) RBD-binding B cells (P=0.1448, Fig. 2e) and a significantly reduced frequency of antigen-experienced Spike-binding B cells compared to normal weight controls 6 months after second vaccine dose (P=0.038, Fig. 2f). Conversely, antigen-specific T cell responses quantified by ELISpot were comparable in people with severe obesity and normal weight controls (Fig. S2f in the Extended Data).

### Response to booster vaccination

We next studied the response to a third (booster) dose of mRNA vaccine (BNT162b2 or mRNA1273) in people with severe obesity (n=28) and normal weight controls (n=16). As expected, levels of anti-Spike and anti-RBD IgG antibodies increased markedly at Day 28 (Fig. 3a and Fig. S3a in the Extended Data). Surprisingly, peak levels were higher in people with severe obesity than normal weight controls (P=0.0052, Fig. 3a and P=0.0014, Fig. S3a in the Extended Data). This finding argues against vaccine delivery failure in people with obesity due to short needle length^21^ (longer needles are recommended for people with BMI>40kg/m^2^ in the UK) and indicates that a fixed rather than weight-adjusted dosing schedule is appropriate for Covid-19 vaccination. Neutralizing antibody titres were nonetheless comparable between the two groups at Day 28 (Fig. 3b). This suggests that, for a given level of antibodies, there was an overall reduction in neutralizing capacity in people with obesity. Elevated anti-RBD levels suggest that severe obesity does not lead to a failure to target neutralizing Spike epitopes, but rather that the impairment in neutralizing capacity may result from a lack of high-affinity antibodies. All participants generated an NT50 > 100, and 61% (n=17) of participants with severe obesity and 67% (n=8) of normal weight controls generated an NT50 > 1,000.

**Figure 3.**
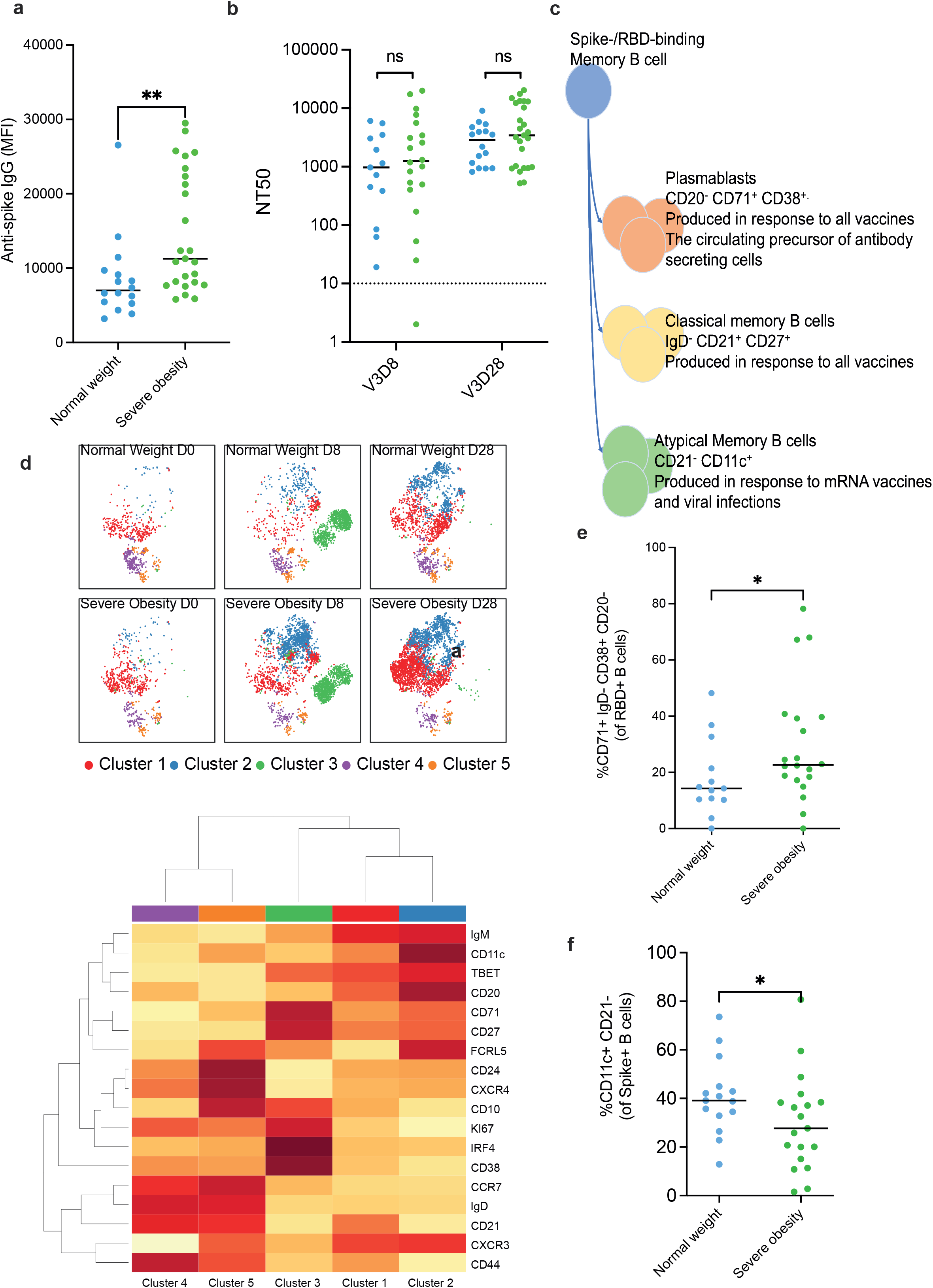
Immune response to third (booster) dose SARS-CoV-2 vaccination. People with severe obesity (green) and normal weight controls (blue) were studied at Day 8 (D8) and Day 28 (D28) after the third vaccine dose (V3). **a,** shows peak levels (Day 28) of anti-Spike IgG antibodies (MFI, mean fluorescence intensity). **b,** depicts neutralizing titres at 50% inhibition (NT50) against wild type SARS-CoV-2 at days 8 and 28, with the dotted line indicating the limit of quantitation. **c,** is a schematic depicting the differentiation of B cells in response to vaccine administration. **d,** shows the results from high dimensional spectral flow cytometry to enumerate and phenotype SARS-CoV-2 Receptor Binding Domain binding (RBD+) and Spike binding (Spike+) B cells. Unsupervised tSNE (t-distributed stochastic neighbour embedding) analysis of B cells at different time points was performed (Methods). This showed that B cells cluster into groups, identified by cell surface and intracellular proteins annotated on the heat map (lower panel). **e,** depicts the frequency of antibody secreting cells (IgD- CD71+ CD38+ CD20) 8 days after third dose vaccination and **f,** the frequency of Spike-binding (Spike+) atypical memory (CD21-CD11c+) B cells 28 days after the third vaccine dose. Panels **a, b, e, f**; horizontal bars indicate the median; ns, not significant, *P<0.05, **P<0.01.

To better understand the specific impairment in humoral immunity associated with severe obesity, we next used high dimensional spectral flow cytometry to enumerate and phenotype SARS-CoV-2 RBD and Spike binding B cells (Fig. 3c and Figs. S3c-e in the Extended Data). Unsupervised tSNE analysis (t-distributed stochastic neighbour embedding) of RBD-binding (RBD+) B cells showed an expansion of antigen-specific B cells with an antibody secreting cell phenotype (IgD- CD71+ CD38+ CD20) 8 days after the third vaccine dose (Fig. 3d), which was greater in individuals with severe obesity than normal weight controls (Fig. 3e) and consistent with their increased antibody levels. In addition, 28 days after the third vaccine dose, individuals with severe obesity had a lower frequency of Spike-binding atypical memory (CD21- CD11c+) B cells (Fig. 3f), suggesting that the humoral immune response elicited in people with severe obesity differs from that seen in normal weight individuals (Figs. S4a-c in the Extended Data). Interestingly, the number of circulating T follicular helper (Tfh) cells, a circulating biomarker of the germinal centre reaction^26^, did not differ between groups (Figs. S4d-e in the Extended Data), suggesting that the enhanced expansion of plasmablasts represents the rapid recall of memory B cells to become antibody secreting cells independent of Tfh cell help. Consistent with the absence of changes in Tfh cells, antigen-specific T cell responses quantified by ELISpot, and the number of regulatory T cells were comparable in people with severe obesity and normal weight controls (Fig. S2b, Figs. S4f-g in the Extended Data).

### Waning of humoral immunity after booster vaccination

The lower neutralizing antibody titres observed in people with severe obesity prior to booster vaccination could reflect a reduction in either the peak response to primary vaccination, or its longevity. We therefore measured longitudinal antibody levels for 105 days (15 weeks) after the third dose of vaccine. We found more rapid waning of anti-Spike and anti-RBD IgG levels and neutralizing antibody titres in individuals with severe obesity (P=0.0057 for percentage change in anti-Spike IgG; P=0.0087 for percentage change in anti-RBD IgG; P=0.0220 for percentage change in NT50; Fig. 4a-c and Figs. S5a-b in the Extended Data). A similar trend was observed in neutralizing capacity against the Omicron variant of SARS-CoV-2 (BA.1) (Fig. S5c in the Extended Data). Conversely, antigen-specific T cell responses quantified by ELISpot remained comparable in people with severe obesity and normal weight controls at day 105 (Fig. 4d). Taken together, these data indicate that severe obesity leads to a failure in the maintenance of humoral immunity following SARS-CoV-2 vaccination, associated with an increased risk of severe Covid-19.

**Figure 4:**
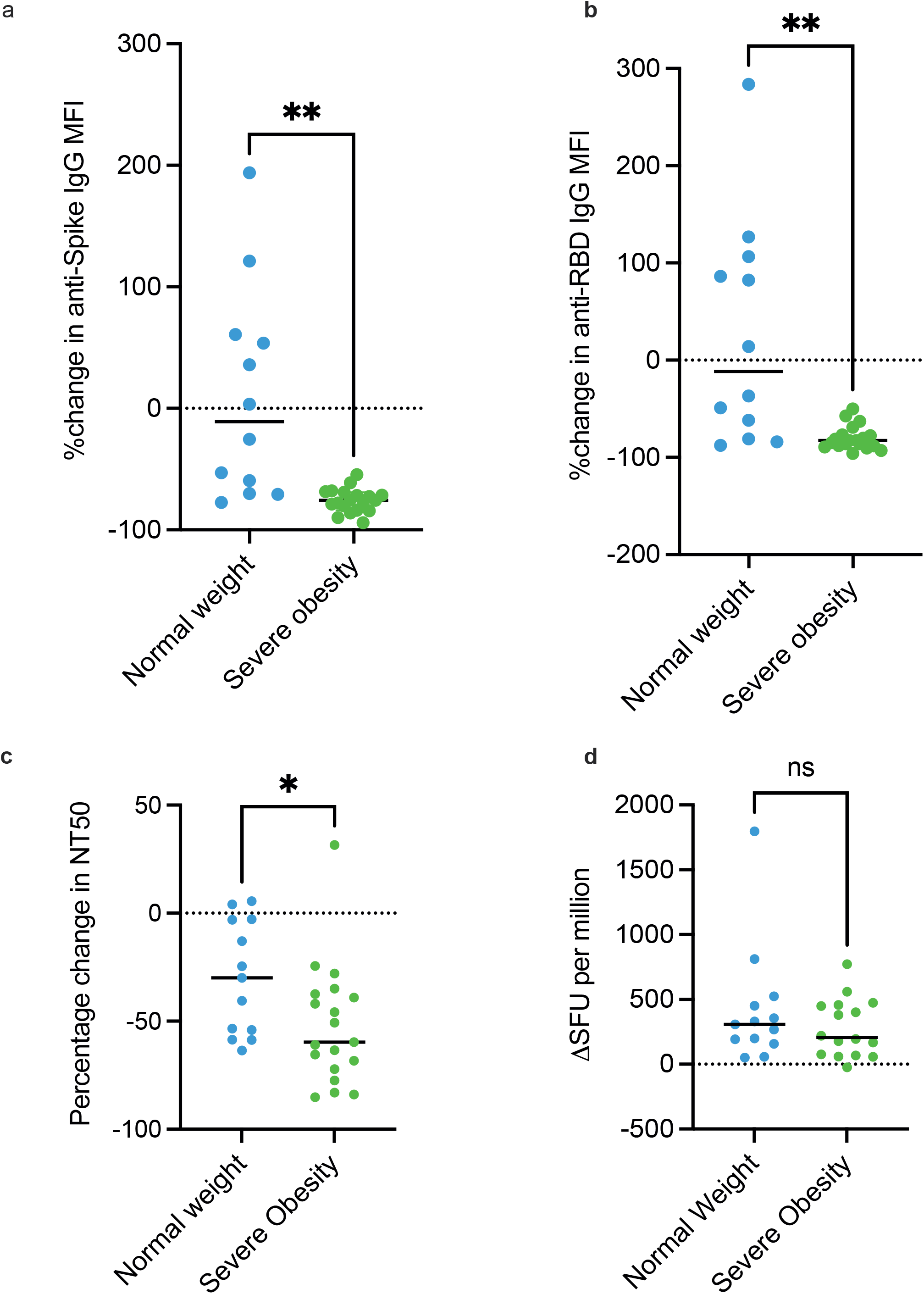
Third dose SARS-CoV-2 vaccine-induced immunity in people with severe obesity. People with severe obesity (green) and normal weight controls (blue) were studied at day 28 and day 105 after the third vaccine dose. **a,** shows the percentage (%) change in anti-Spike IgG antibody levels (MFI, mean fluorescence intensity) and **b,** the % change in anti-Receptor Binding Domain (RBD) IgG antibody levels between these two time-points. **c,** shows the % change in neutralizing titres at 50% inhibition (NT50) against wild type SARS-CoV-2, with the dotted line indicating the limit of quantitation. **d,** shows T cell responses quantified by Elispot. SFU = Interferon gamma spot forming units. Panels **a, b, c, d;** horizontal bar indicates the median. Participants who reported a positive SARS-CoV-2 RT-PCR test between day 28 and 105 or who had positive anti-nucleocapsid antibodies at day 105 were excluded from these analyses. ns = non-significant, *P<0.05, **P<0.01.

## Discussion

Obesity is an established risk factor for severe Covid-19^2^. Here, we show that this risk persists even after SARS-CoV-2 vaccination and increases with time after the primary vaccination course. Our studies of over 0.5 million vaccinated people with obesity and >98,000 vaccinated people with severe obesity are consistent with increased BMI leading to a reduction in the maintenance of vaccine-induced immunity and increased breakthrough infections. In keeping with this, we find evidence of reduced neutralizing antibody capacity 6 months after primary vaccination in individuals with severe obesity. These changes in antibody kinetics are associated with altered B cell differentiation, and a dissociation between anti-Spike antibody levels and neutralizing capacity. A similar, relative reduction in neutralizing capacity has previously been observed in patients with severe Covid-19 in other settings^27,28^ and may reflect either lower antibody affinities, or differential targeting of neutralizing and non-neutralizing Spike epitopes.

Our findings in people with severe obesity are consistent with previous studies showing that the acute immune response to SARS-CoV-2 vaccines is comparable in people with obesity and normal weight people^15–20^. Some of these studies suggested that the duration of vaccine-induced immunity may be reduced in people with obesity^16–18^. These studies all relied on measurements of immunity at a single time-point and used different assays/endpoints (e.g. self-reported home antibody tests^19^ or an assumption of effectiveness in those who tested negative for Covid-19 by RT-PCR^20^). Here, by prospectively measuring B cell and T cell responses as well as neutralizing capacity of antibodies to authentic virus in vaccinated people with severe obesity and normal weight controls over time, we demonstrate that the waning of humoral immunity associated with SARS-CoV-2 vaccines^29^ is accelerated in people with severe obesity.

Our data suggest that obesity may lead to a short-lived antibody response. Antibody production after vaccination can occur via two pathways, which differ in terms of quality or longevity^30^. The first pathway, the extrafollicular response, results in an initial burst of antibodies early after vaccination. This response is short-lived, with no additional diversification of the B cell repertoire, and its contribution to long-term immunity is therefore minimal. The second pathway, the germinal centre (GC) reaction, produces memory B cells and long-lived antibody secreting plasma cells that can persist long-term in the bone marrow. After third dose/booster vaccination, we found people with severe obesity had an increased number of circulating antibody secreting cells which are biomarkers of the extrafollicular pathway, but not circulating Tfh cells which are biomarkers of the germinal centre response^26^. We therefore hypothesize that in obesity, the extrafollicular pathway is preferred at the expense of the GC pathway, although the mechanism that drives this is not known.

Several studies in mice and humans have suggested mechanisms by which obesity may impact on immunity and more specifically on vaccine response^31,32^. Potential contributory factors include dietary constituents such as fatty acids, the infiltration of bone marrow and the spleen by adipocytes and adipose tissue-derived cytokines such as leptin and interleukin-6^33,34^, which modulate the function of CD4+ T Follicular Helper and B lymphocytes critical for the GC^35,36^. Obesity is often characterized as a pro-inflammatory state due to the activation of immune cells (particularly macrophages) in adipose tissue^37,38^. While it has been suggested that the persistence of a chronic pro-inflammatory state in people with obesity may affect their response to immunological challenges such as vaccination, the mechanisms by which such effects may be mediated require further exploration.

Since people with obesity show a reduction in the maintenance of humoral vaccine responses, additional or more frequent booster doses are likely to be required to maintain protection against Covid-19. Because of the high prevalence of obesity^39^, this poses a major challenge for health services and vaccine programs around the world.

**Table 1.**
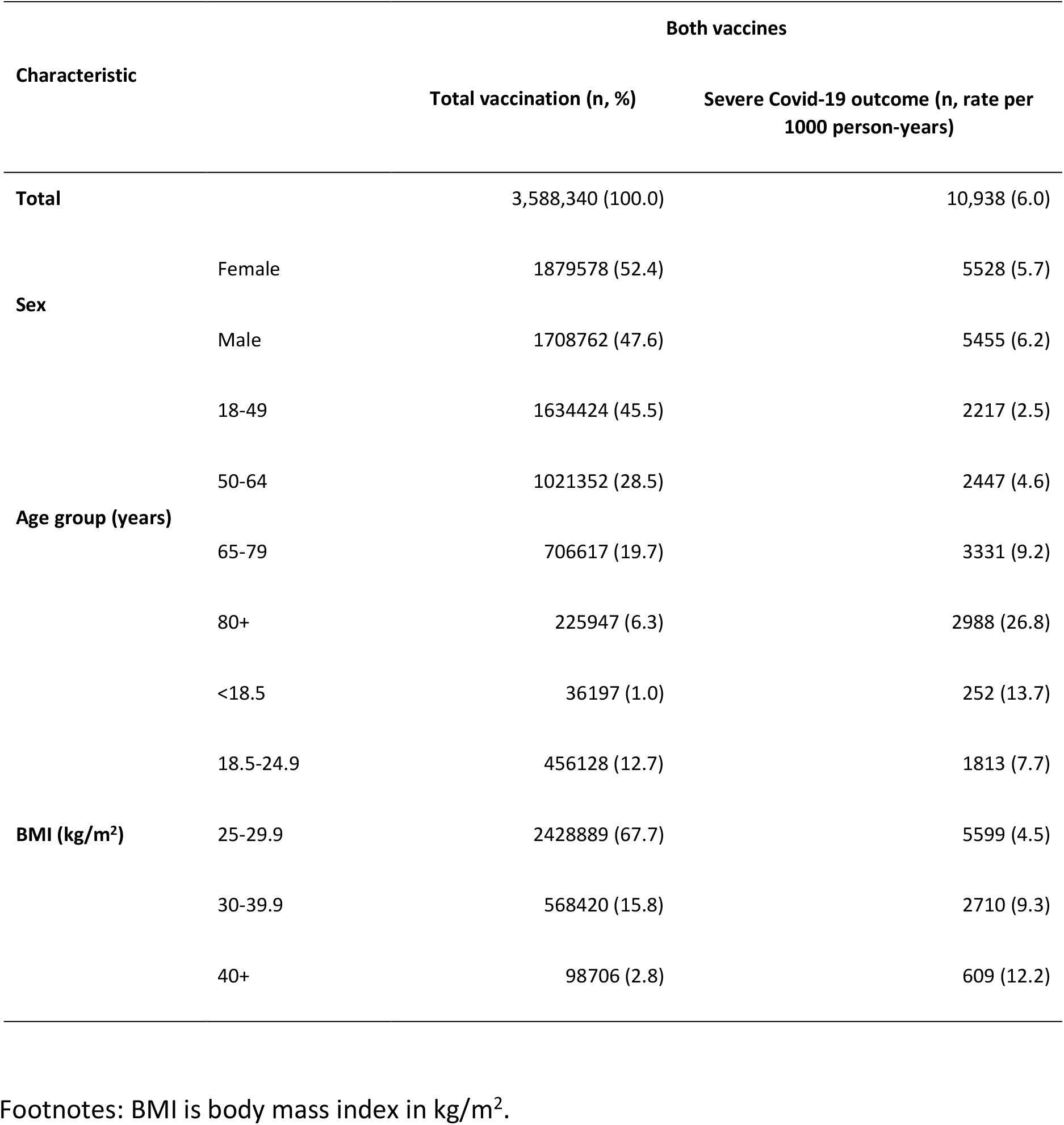
Population characteristics of individuals from EAVE-II who received second and third doses of a SARS-CoV-2 vaccine

## Data Availability

All data produced in the present study are available upon reasonable request to the authors

## ACKNOWLEDGEMENTS

The epidemiological study is part of the EAVE II project. EAVE II is funded by the MRC (MC_PC_19075) with the support of BREATHE—The Health Data Research Hub for Respiratory Health (MC_PC_19004), which is funded through the UK Research and Innovation Industrial Strategy Challenge Fund and delivered through the Health Data Research UK. This research is part of the Data and Connectivity National Core Study, led by Health Data Research UK in partnership with the Office for National Statistics and funded by UK Research and Innovation (grant ref MC_PC_20058) and the National Core Studies - Immunity. Additional support has been provided through Public Health Scotland, the Scottish Government Director-General Health and Social Care and the University of Edinburgh. The original EAVE project was funded by the National Institute for Health Research (NIHR) Health Technology Assessment programme (11/46/23). We thank Dave Kelly from Albasoft (Inverness, UK) for his support with making primary care data available, and Wendy Inglis-Humphrey, Vicky Hammersley, and Laura Brook (University of Edinburgh, Edinburgh, UK) for their support with project management and administration.

The SCORPIO study was supported by the Medical Research Council (MR/W020564/1, a core award to J.E.T.; MC_UU_0025/12 and MR/T032413/1, an award to N.J.M.) and the Medical Research Foundation (MRF-057-0002-RG-THAV-C0798). Additional support was provided by NHSBT (WPA15-02 to N.J.M.), Addenbrooke’s Charitable Trust (900239 to N.J.M.) and the National Institute for Health Research (NIHR) Cambridge Biomedical Research Centre (N.J.M. and I.S.F.) and NIHR BioResource. M.A.L is supported by the Biotechnology and Biological Sciences Research Council (BBS/E/B/000C0427, BBS/E/B/000C0428), and is a Lister Institute Fellow and an EMBO Young Investigator. I.M.H. is supported by a CIMR PhD studentship; H.J.S. by a Sir Henry Dale Fellowship jointly funded by Wellcome and the Royal Society [109407] and a BBSRC institutional programme grant [BBS/E/B/000C0433]. I.S.F. is supported by Wellcome (207462/Z/17/Z), Botnar Fondation, the Bernard Wolfe Health Neuroscience Endowment and a NIHR Senior Investigator Award. The PITCH study was funded by the UK Department of Health and Social Care. S.J.D. is funded by an NIHR Global Research Professorship (NIHR300791). P.K. is an NIHR Senior Investigator and is funded by Wellcome (WT109965MA). We thank the staff of the obesity clinic at Cambridge University Hospitals for their support in recruitment of participants to the SCORPIO study. Clinical studies were performed in the Wellcome-MRC IMS Translational Research Facility (TRF) supported by a Wellcome Major Award [208363/Z/17/Z]) and in the Cambridge NIHR Clinical Research Facility. WHO International Standard 20/136 was a kind gift from Heli Harvala and David Roberts (NHSBT). The funding bodies had no role in the design or conduct of the study; collection, management, analysis or interpretation of the data; preparation, review or approval of the manuscript or the decision to submit the manuscript for publication. The views expressed are those of the authors and not necessarily those of the NIHR, the Department of Health and Social Care, or the UK government.

## Author contributions

AAvdK, JEDT and ISF designed and led the study. UA, CM, CR and AS performed the analyses in the EAVE II cohort; AS is the guarantor of this work. AAvdK, SS, MS, BV, EH, JS, AJP, HS, NK, FR and ISF recruited the SCORPIO cohort, performed clinical studies and/or analysed clinical data. ECH co-ordinated laboratory studies including extraction of lymphocytes from clinical samples and T cell assays with SS, LHB, TEM, MJOR, TPG, LPG, MAR, AF; these experiments were overseen by JEDT. NJM provided oversight of studies of neutralizing antibodies with WT and Omicron variants; experiments were performed by PPG with IDR, JAS. RD, SE and LCG performed measurements of anti-S, anti-N antibodies. MAL led experiments of quantitation of lymphocyte types and subsets by flow cytometry including computational analysis (tSNE); these experiments were performed by WSF, SMG, HJS, IMH, ST and SI. BK, SJD, PK and the PITCH Consortium contributed samples from normal weight controls recruited to the PITCH study. All authors approved the final version of the manuscript.

## Competing interest declaration

AS is a member of the Scottish Government’s Standing Committee on Pandemic Preparedness and the Risk Stratification Subgroup of the UK Government’s New and Emerging Respiratory Virus Threats Advisory Group (NERVTAG). He was a member of AstraZeneca’s Thrombotic Thrombocytopenic Task Force. All roles are unremunerated. SJD is a Scientific Advisor to the Scottish Parliament on COVID-19 for which she receives a fee. All other authors have no conflict of interest to declare.

## METHODS

### ETHICAL APPROVAL AND STUDY POPULATIONS

#### EAVE II study

The EAVE II surveillance platform drew on near real-time nationwide health care data for 5.4 million individuals (∼99%) in Scotland^1–4^. It includes information on clinical and demographic characteristics of each individual, their vaccination status and type of vaccine used and information on positive SARS-COV-2 infection and subsequent hospitalization or death. Ethical approval was granted by the National Research Ethics Service Committee, Southeast Scotland 02 (reference number: 12/SS/0201) for the study using the Early Pandemic Evaluation and Enhanced Surveillance of Covid-19 (EAVE II) platform. Approval for data linkage was granted by the Public Benefit and Privacy Panel for Health and Social Care (reference number: 1920-0279). Individual written patient consent was not required for this analysis.

Using data from the EAVE II platform, we examined the impact of obesity (using BMI measurements), clinical and demographic characteristics including time since receiving the second and third vaccine dose, previous history of testing positive for Covid-19, gap between vaccine doses and dominant variant in the background, of fully vaccinated adults in Scotland who experienced severe Covid-19 outcomes. The cohort analyzed for this study consisted of individuals aged 18 and over who were administered with at least two doses of BNT162b2 mRNA, ChAdOx1 nCoV-19 or mRNA-1273 vaccines between December 8, 2020 and March 19, 2022. Follow-up began 14 days after receiving the second dose until Covid-19 related hospitalization, Covid-19 related death or the end of study period (i.e., March 19, 2022). All the Covid-19 related hospital admissions or deaths were selected between September 14, 2020 and March 19, 2022. Patients without immunosuppression had their primary vaccination schedule with two doses and so the third dose is a booster. For people with immunosuppression, the primary vaccination schedule was for three vaccine doses. BMI was available for individuals based on last recorded measurement within their primary care record. Where the BMI was missing, it was imputed using ordinary least squares regression with all other independent variables included as predictors.

##### Definition of outcomes

The primary outcome of interest was severe Covid-19, which was defined as Covid-19 related hospital admission or death, 14 days or more after receiving the second vaccine or booster dose^5^. Covid-19 related hospital admission was defined as hospital admission within 14 days of a positive reverse-transcriptase polymerase chain reaction (RT-PCR) test or Covid-19 as reason for admission or a positive SARS-CoV-2 RT-PCR test result during an admission where Covid-19 was not the reason for admission. Covid-19 related mortality was defined as either death for any reason within 28 days of a positive RT-PCR test or where Covid-19 was recorded as the primary reason for death on the death certificate.

##### Population characteristics and confounders

Characteristics of interest were defined at baseline on December 8, 2020 and included age, sex, socioeconomic status based on quintiles of Scottish Index of Multiple Deprivation (SIMD), urban or rural place of residence, BMI, previous natural infection from SARS-CoV-2 prior to second dose of the vaccine (classified as 0-3 months, 3-6 months, 6-9 months, ≥9 months prior to second infection), number of pre-existing comorbidities known to be linked with severe Covid-19 outcome and being in a high risk occupational group defined as someone with undergoing regular RT-PCR testing^2^. BMI was grouped as <18.5 (underweight), 18.5-24.9 (normal weight), 25-29.9 (overweight), 30-39.9 (obese) and >=40 kg/m^2^ (severely obese) according to World Health Organization (WHO) criteria.

##### Statistical analysis EAVE II

We calculated the frequency and rate per 1,000 person-years of severe Covid-19 outcomes for all demographic and clinical factors. Generalized Linear Models (GLM) assuming a Poisson distribution with person-time as an offset representing the time at risk were used to derive rate ratios (RRs) with 95% confidence intervals (CIs) for the association between demographic and clinical factors and Covid-19 related hospitalization or death. Adjusted rate ratios (aRRs) were estimated adjusting for all confounders including age, sex, SIMD, time since receiving the second dose of vaccine, pre-existing comorbidities, the gap between vaccine doses, previous history of SARS-CoV-2 infection and calendar time. R (version 3.6.1) was used to carry out all statistical analyses.

#### SARS-CoV2 vaccination response in obesity (SCORPIO) clinical study

Clinical studies in people with severe obesity and normal weight controls were approved by the National Research Ethics Committee and Health Research Authority (East of England – Cambridge Research Ethics Committee (SCORPIO study, SARS-CoV-2 vaccination response in obesity amendment of ‘‘NIHR BioResource’’ 17/EE/0025)). Each subject provided written informed consent. All studies were conducted in accordance with the Declaration of Helsinki.

People with severe obesity (class II/ III WHO criteria of BMI ≥ 40 kg/m^2^ or BMI ≥ 35 kg/m^2^ with obesity-associated medical conditions such as type 2 diabetes, hypertension) who attended the obesity clinic at Cambridge University Hospitals NHS Trust and had received two doses of SARS-CoV-2 vaccination (first and second dose of ChAdOX1 nCoV-19 or BNT162b2 mRNA) between December 2021 and May 2022, were invited to take part. People with acquired (HIV, immunosuppressant drugs) or congenital immune deficiencies and cancer were excluded. Third dose vaccinations (BNT162b2, Pfizer BioNTech or half dose mRNA1273, Moderna) were administered as part of the NHS vaccination program. UK Health Security Agency policy recommends the use of longer needles (38 vs 25 mm) in people with severe obesity.

Additional normal weight controls were recruited in Oxford, UK as part of the PITCH study under the GI Biobank Study 16/YH/0247, approved by Yorkshire & Humber Sheffield Research Ethics Committee, which was amended for this purpose on 8 June 2020. Samples obtained 6 months after primary course were included.

Clinical and immunological measurements were taken before the booster vaccination, 8 (−3) days, 28 (+-7) days and 105 (+-7) days after vaccination. Third dose vaccinations were administered as part of the NHS vaccination program and where mRNA vaccines (BNT162b2 or mRNA1273 (Moderna). Clinical data regarding co-morbidities associated with obesity was obtained from the medical records. Supplementary Table 3 details the demographic characteristics of this cohort. Healthy healthcare workers were enrolled into the longitudinal OPTIC cohort in Oxford, UK between May 2020 and May 2021 as part of the PITCH consortium. PITCH participants were sampled between July and November 2021, a median of 185 days (range 155-223) days after receiving a second vaccination with ChAdOX1 nCoV-19 or BNT162b2 mRNA vaccine. All PITCH participants were classified as infection-naïve, as defined by never having received a positive lateral flow or PCR test for SARS-CoV-2, and negative anti-nucleocapsid antibodies at the time of their first vaccination. Therefore, a total of 28 people with severe obesity and 41 normal weight controls were evaluated 6 months after primary course of vaccination, whereas for the response to third dose vaccination, 16 normal weight controls were studied.

Of the 28 recruited people with severe obesity, 2 had positive anti-N antibodies and reported a positive PCR test before their third dose vaccination. They were excluded from further analysis. In addition, between day 28 and 105, 2 people with severe obesity reported positive SARS-CoV-2-tests (lateral flow test or PCR tests as per UK guidelines at the time). They were excluded from the day 28 to day 105 analysis. In addition, one of the normal weight participants had positive anti-N antibodies who had not had a PCR test, before their third dose vaccination. This participant was excluded from the before and after third dose analysis. In addition, between day 28 and 105, 2 normal weight people reported positive SARS-CoV-2-tests (lateral flow test or PCR tests as per UK guidelines at the time, one of those participant on two separate occasions). They have been excluded from the day 28 to day 105 analysis.

Peripheral blood samples were acquired in either lithium heparin (PBMCs) or serum separating tubes. Serum tubes were centrifuged at 1600 x g for 10 minutes at room temperature (RT) to separate serum from the cell pellet before being aliquoted and stored at −80°C until use. Peripheral blood mononuclear cells (PBMCs) were isolated by layering over lymphoprep density gradient medium (Stemcell Technologies) followed by density gradient centrifugation at 800 x g for 20 minutes at RT. PBMCs were isolated and washed twice using wash buffer (1X PBS, 1% foetal calf serum, 2mM EDTA) at 400 x g for 10 minutes at 4°C. Isolated PBMCs were resuspended in freezing media, aliquoted and stored at −80°C for up to a week before being transferred to liquid nitrogen until use.

#### SARS-CoV-2 serology by multiplex particle-based flow cytometry (Luminex)

Recombinant SARS-CoV-2 anti-nucleocapsid (N), anti-Spike (S) and anti-Receptor-Binding Domain (RBD) antibodies were measured by multiplex particle-based flow cytometry (Luminex)^6,7^. Recombinant SARS-CoV-2 nucleocapsid (N), spike (S) and receptor-binding domain (RBD) proteins were covalently coupled to distinct carboxylated bead sets (Luminex; Netherlands), forming a 3-plex assay. The S protein construct used is described by Xiong et al^8^, and the RBD construct was described by Stadlbauer et al^9^. The N protein used is a truncated construct of the SARS-CoV-2 N protein comprising residues 48–365 (both ordered domains with the native linker) with an N terminal uncleavable hexahistidine tag. N was expressed in *E*. *Coli* using autoinducing media for 7h at 37°C and purified using immobilised metal affinity chromatography (IMAC), size exclusion and heparin chromatography. Beads were first activated with 1-ethyl-3-[3-dimethylaminopropyl]carbodiimide hydrochloride (Thermo Fisher Scientific) in the presence of N-hydroxysuccinimide (Thermo Fisher Scientific), according to the manufacturer’s instructions, to form amine-reactive intermediates. The activated bead sets were incubated with the corresponding proteins at a concentration of 50 μg/ml in the reaction mixture for 3 h at room temperature on a rotator. Beads were washed and stored in a blocking buffer (10 mM PBS, 1% BSA, 0.05% NaN3). The N-, S- and RBD-coupled bead sets were incubated with patient sera at 3 dilutions (1/100, 1/1000, 1/10000) for 1 h in 96-well filter plates (MultiScreen HTS; Millipore) at room temperature in the dark on a horizontal shaker. Fluids were aspirated with a vacuum manifold and beads were washed three times with 10 mM PBS/0.05% Tween 20. Beads were incubated for 30 min with a PE-labeled anti–human IgG-Fc antibody (Leinco/Biotrend), washed as described above, and resuspended in 100 μl PBS/ Tween. They were then analysed on a Luminex analyser (Luminex / R&D Systems) using Exponent Software V31. Specific binding was reported as mean fluorescence intensity (MFI).

#### Neutralizing antibodies to SARS-CoV-2

Neutralization of authentic SARS-CoV-2 virus was measured using reporter cells expressing a protease-activatable luminescent biosensor^10^. The SARS-CoV-2 viruses used in this study were a wildtype (lineage B) isolate (SARS-CoV-2/human/Liverpool/REMRQ0001/2020), a kind gift from Ian Goodfellow (University of Cambridge), isolated by Lance Turtle (University of Liverpool), David Matthews and Andrew Davidson (University of Bristol) ^11,12^, and an Omicron (lineage B.1.1.529) variant, a kind gift from Ravindra Gupta^13^.

Sera were heat-inactivated at 56°C for 30 mins before use, and neutralizing antibody titres at 50% inhibition (NT50s) measured as previously described^10,14^. In brief, luminescent HEK293T-ACE2-30F-PLP2 reporter cells (clone B7) expressing SARS-CoV-2 Papain-like protease-activatable circularly permuted firefly luciferase (FFluc) were seeded in flat-bottomed 96-well plates. The next day, SARS-CoV-2 viral stock (MOI=0.01) was pre-incubated with a 3-fold dilution series of each serum for 2 h at 37°C, then added to the cells. 16 h post-infection, cells were lysed in Bright-Glo Luciferase Buffer (Promega) diluted 1:1 with PBS and 1% NP-40, and FFluc activity measured by luminometry. Experiments were conducted in duplicate.

To obtain NT50s, titration curves were plotted as FFluc vs log (serum dilution), then analysed by non-linear regression using the Sigmoidal, 4PL, X is log(concentration) function in GraphPad Prism. NT50s were reported when (1) at least 50% inhibition was observed at the lowest serum dilution tested (1:10), and (2) a sigmoidal curve with a good fit was generated. For purposes of visualisation and ranking, samples with no neutralizing activity were assigned an arbitrary NT50 of 2. Samples for which visual inspection of the titration curve indicated inhibition at low dilutions, but which did not meet criteria (1) and (2) above, were assigned an arbitrary NT50 of 4. Unless otherwise indicated, all NT50s shown refer to neutralization of wildtype virus.

To enable comparison with other studies, the neutralizing capacity of World Health Organisation International Standard 20/136 (WHO IS 20/136) against wildtype SARS-CoV-2 was measured in 5 independent experiments, yielding a geometric mean NT50 of 1967 (Fig. S5 in extended data). This standard comprises pooled convalescent plasma obtained from 11 individuals which, when reconstituted, is assigned an arbitrary neutralizing capacity of 1000 IU/ml against early 2020 SARS-CoV-2 isolates^15^. NT50s against wildtype SARS-CoV-2 from this study may therefore be converted to IU/ml using a calibration factor of 1000/1967 (0.51), with a limit of quantitation of 5.1 IU/ml (corresponding to an NT50 of 10).

#### T cell cytokine production

T cell responses were assessed using an ELISpot assay^16^. T cell ELISpot assays were performed using the PITCH Standard Operating Procedure ^16^. Cryopreserved PBMCs were thawed using RMPI media supplemented with 1% (v/v) Penicillin/Streptomycin (Sigma) containing 0.01% (v/v) Benzoase Nuclease (Merck), and then rested for 2-3 hours in RPMI media supplemented with 10% (v/v) Human AB Serum (Sigma) and 1% (v/v) Penicillin/Streptomycin (Sigma) at 37°C. Plates precoated with capture antibody (Mabtech, mAb 1-D1K) were washed three times with sterile phosphate buffered saline (PBS) and then blocked with RPMI media supplemented with 10% (v/v) Human AB Serum and 1% (v/v) Penicillin/Streptomycin at 37°C for 1-2 hours. Overlapping peptide pools (18-mers with 10 amino acid overlap. Mimotopes) representing the spike (S), Membrane (M) or nucleocapsid (N) SARS-CoV-2 proteins were added to 200,000 PBMCs/well at a final concentration of 2µg/ml. Pools consisting of CMV, EBV, influenza, and Tetanus toxoid peptides at a final concentration of 2µg/ml (CEFT; Proimmune) and Concanavalin A (Sigma) were used as positive controls. DMSO (Sigma) was used as the negative control at the equivalent concentration to the peptides. Plates were incubated at 37°C, 5% humidity, for 18 hours. Wells were washed with PBS 0.05% (v/v) Tween (Sigma) seven times before incubation for 2 hours at room temperature with the ELISpot PLUS kit biotinylated detection antibody (clone 7-B6-1) diluted in PBS to 1 µg/ml, 50µl per well. Wells were washed with PBS with 0.05% (v/v) Tween, and then incubated with the ELISpot PLUS kit streptavidin-ALP, diluted in PBS to 1µg/ml for 1 hour at room temperature. Wells were washed with PBS 0.05% (v/v) Tween and colour development was carried out using 1-step NBT/BCIP Substrate Solution. 50µl of filtered NBT/BCIP was added to each well for 5 minutes at RT. Colour development was stopped by washing the plates with cold tap water. Plates were left to air dry for 48 hours and the scanned and analysed using the AID iSpot Spectrum ELISpot reader (software version 7.0, Autoimmune Diagnostika GmbH, Germany). The average spot count of the control wells were subtracted from the test wells for each sample to quantify the antigen-specific responses. Results are expressed as spot forming units (SFU) per 10^6^ PBMCs. Analysis was completed using GraphPad Prism software version 9.3.1. The comparison of means between groups was performed using two-way, mixed model ANOVA.

#### Quantitation of lymphocyte types and subsets by flow cytometry

##### Generation of fluorescent RBD-specific and Spike-specific B cell probes

BirA and RBD-avi-His plasmids were subcloned in DH5α Competent Cells (Invitrogen™, Cat# 18265017) and purified using EndoFree Plasmid Mega Kit (Qiagen, Cat# 12381) following manufacturer’s protocols. Then, biotinylated SARS-CoV-2 RBD with C-terminal Avi and hexahistidine tags was expressed by transient co-transfection transfection of RBD-avi-His and BirA expression plasmids in HEK-293F cells (ThermoFisherScientific Cat# R79007) using PEI MAX (Polysciences Cat# 24765). 1.2 L of culture at a density of 1.0 x 10^6^ cells/mL was supplemented with 175 μM biotin (Sigma-Aldrich Cat# B4501) and transfected with 600 µg total DNA at a 4:1 ratio (RBD:BirA) using 12 ml PEI MAX (1 mg/mL). Culture medium was harvested 6 days post-transfection and protein purified using Ni-NTA agarose beads (Qiagen Cat# 30210). Eluted protein was further purified by size-exclusion chromatography on a HiLoad Superdex 200 pg 16/600 column (Cytiva Cat# 28989335) equilibrated in 1X PBS. Peak fractions were pooled, concentrated in a 10 kDa MWCO centrifugal filter (Merck Cat# UFC801024) and snap frozen in liquid nitrogen.

BirA and Spike-avi-His plasmids were subcloned in DH5α Competent Cells (Invitrogen™, Cat# 18265017) and purified using EndoFree Plasmid Mega Kit (Qiagen, Cat# 12381) following manufacturer’s protocols. Biotinylated SARS-CoV-2 S protein with C-terminal Avi and hexahistidine tags was expressed by transient co-transfection of S protein and BirA expression plasmids in HEK-293F cells using PEI MAX. Briefly, 400 ml of culture at a density of 1.0 x 10^6^ cells/ml was supplemented with 175 µM biotin and transfected with 400 µg total DNA at a 4:1 ratio (S-protein:BirA) using 600 µl PEI MAX (1 mg/ml). Culture medium was harvested 6 days post-transfection and protein purified using Ni-NTA agarose beads in batch mode. Beads were loaded into a glass gravity column, washed with 20 ml of PBS with 5 mM imidazole, followed by elution of protein in 5 x 1 ml fractions of PBS with 300 mM imidazole. Eluted protein was further purified by size-exclusion chromatography on a HiLoad Superdex 200 pg 16/600 column equilibrated in 1X PBS. Peak fractions were pooled, concentrated in a 30 kDa MWCO centrifugal filter (Merck Cat# UFC803024) and snap frozen in liquid nitrogen.

Monomers were combined with fluorescently labelled streptavidin (BioLegend) at a 3.95:1 molecular ratio, to ensure complete tetramerization of streptavidin molecules. RBD was added to streptavidin in 10% increments, with a 10-minute interval between each addition, and was gently mixed throughout the tetramerization process. Tetramerization was carried out at room temperature, before storage at 4°C.

##### Spectral flow cytometry

RBD-specific and Spike-specific B cells were measured by high dimension flow cytometry^17^. For flow cytometry stains a single cell suspension was prepared from cryopreserved PBMC samples as follows: 1mL PBMC samples were defrosted in a 37°C water bath, and then immediately diluted into 9mL of pre-warmed RPMI+10% Fetal Bovine serum (FBS). Cells were washed twice with 10 mL of FACS buffer (PBS containing 2% FBS and 1mM EDTA). Cells were then resuspended in 500uL of FACS buffer and cell numbers and viability were determined using a Countess™ automated cell counter (Invitrogen). 5×10^6^ viable cells were transferred to 96-well plates for antibody staining. Cells were then washed once with FACS buffer, and stained with 100 µL of surface antibody mix (including B cell probes) for 2 hours at 4°C. Cells were then washed twice with FACS buffer, and fixed with the eBioscience Foxp3/Transcription Factor Staining Buffer (ThermoFisher #00-5323-00) for 30 min at 4°C. Cells were then washed with 1x eBioscience Foxp3/Transcription Factor Permeabilisation buffer ((ThermoFisher #00-8333-56) twice and stained with intracellular antibody mix in permeabilisation buffer at 4°C overnight. Following overnight staining, samples were washed twice with 1x permeabilisation buffer and once with FACS buffer and acquired on a Cytek^TM^ Aurora. Cells for single colour controls were prepared in the same manner as the fully stained samples. Manual gating of flow cytometry data was done using FlowJo v10.8 software (Tree Star).

#### Statistical analysis SCORPIO study

Analysis was completed using GraphPad Prism software version 9.3.1. The comparison of means or medians between groups was performed using parametric T test or non-parametric Mann-Whitney U test when appropriate. tSNE, FlowSOM and heatmap analysis were performed using R (version 4.1.2) using code that has previously been described ^18^.

---

**Supplementary Figure 1:**
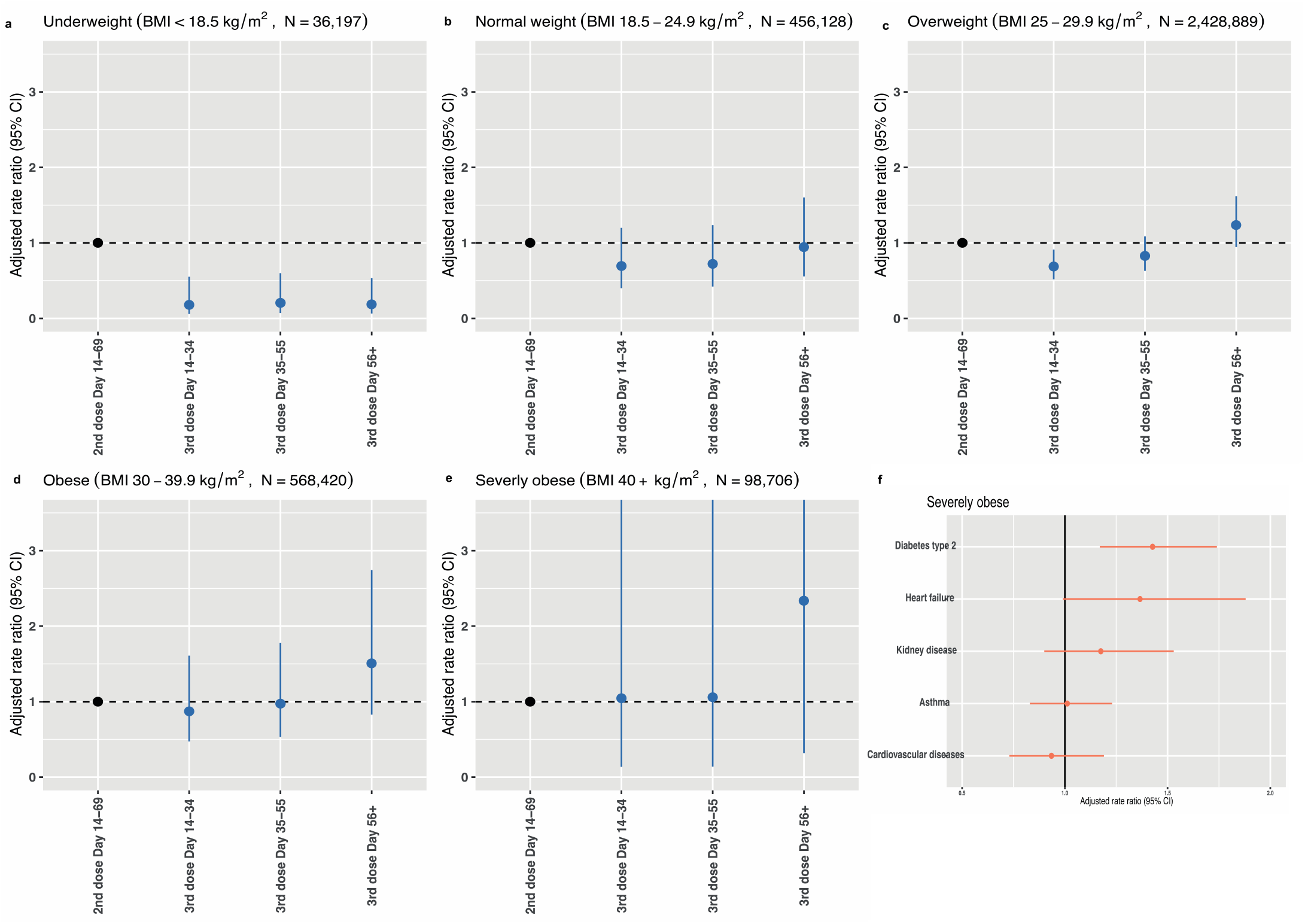
Adjusted rate ratios for hospitalization and death due to Covid-19 in vaccinated people **a-e,** Adjusted rate ratios for hospitalization or death following third (booster) doses per BMI category. Adjusted Rate Ratios (aRR) for hospitalisation or death following third (booster) dose for different body mass index (BMI) categories. Error bars indicate 95% confidence intervals. N indicates number of people in each category. **f**, adjusted rate ratios for hospitalisation and death in severely obese individuals with obesity-associated comorbidities. Error bars indicate 95% confidence intervals.

**Supplementary Figure 2:**
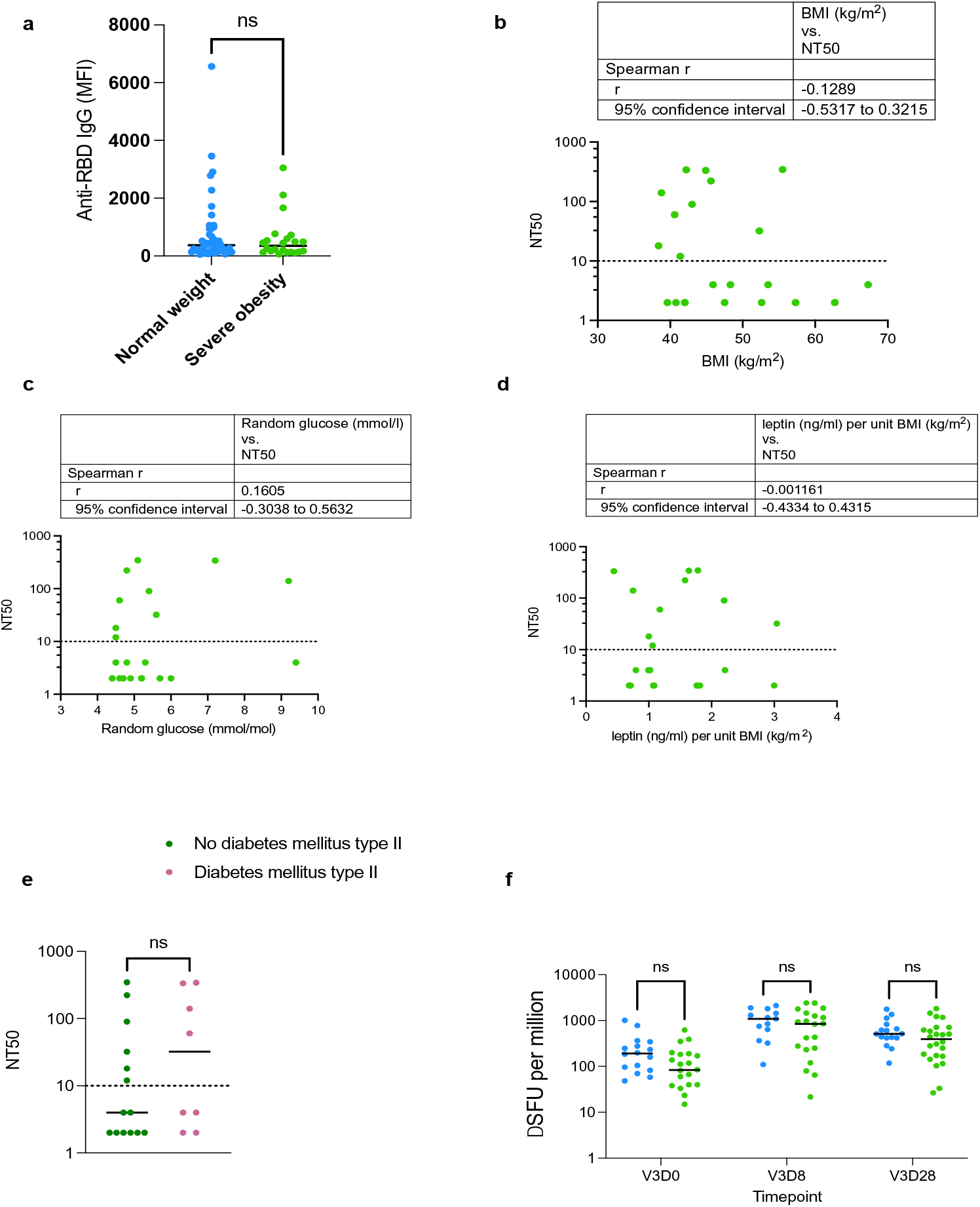
SARS-CoV-2 vaccine-induced immunity in people with severe obesity and normal weight people. **a**, Anti-RBD (receptor binding domain) IgG titres are comparable in people with severe obesity (green) and normal weight individuals (blue) 6 months after primary vaccination course (P=0.5803 in Mann-Whitney test). Each symbol represents an individual person and line indicates the median; ns, not significant. **b-e**, Correlation between body mass index (BMI), random blood glucose, leptin levels per unit BMI and the presence or absence of type II diabetes and neutralizing capacity (NT50) 6 months after second dose. Non-parametric Spearman’s Rho correlations were calculated between NT50 and clinical parameters. Non-parametric Mann-Whitney test was used to compare people with severe obesity without and with diabetes mellitus. Dotted line indicates the limit of quantitation; ns, not significant. **f**, Antigen-specific T cell responses were quantified by ELISpot. Responses were comparable in people with severe obesity (green) and normal weight controls (blue) before and after third dose booster vaccination. Interferon gamma spot forming units (SFU) were quantified. Each symbol represents an individual person and line indicates the median; ns, not significant. V3D0 is before third dose vaccination (V3), V3D8 is 8 days after third dose and V3D28 is 28 days after third dose vaccination.

**Supplementary Figure 3:**
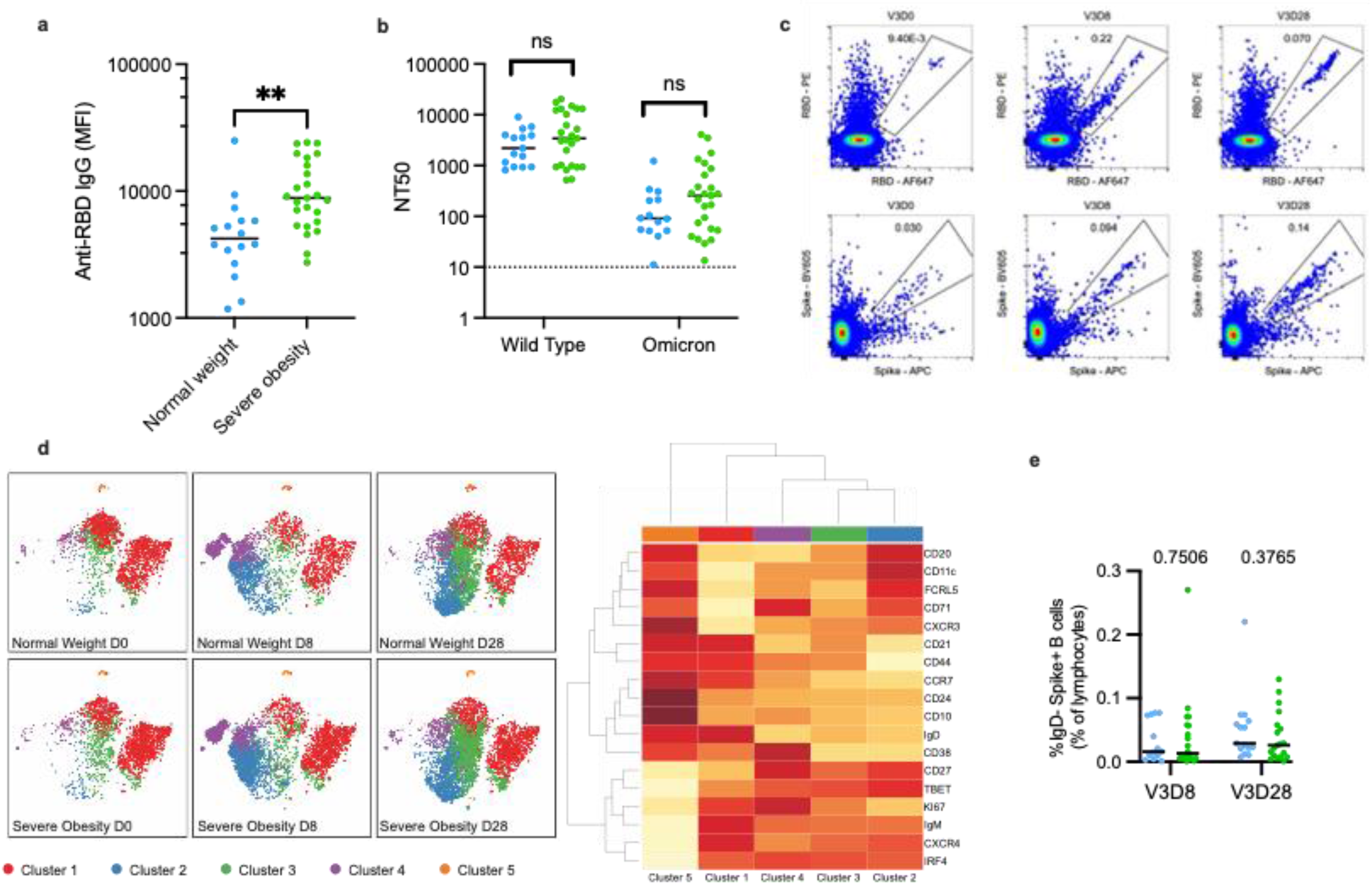
Response to third dose booster vaccination in severe obesity **a**, Anti-RBD (Receptor Binding Domain) IgG levels (MFI, mean fluorescence intensity) were higher in people with severe obesity (green) compared to normal weight controls (blue) (**P=0.0013 in Mann-Whitney test). Each symbol represents an individual person studied at Day 28; line indicates the median. **b**, Neutralizing antibody titres (NT50) against the Omicron variant of SARS-CoV-2 were markedly reduced compared with wild type virus (Day 28), but no difference was observed between groups. Non-parametric Mann-Whitney test was used to compare people with severe obesity (green) and normal weight controls (blue). Individual values are shown, with the dotted line indicating the limit of quantitation; ns, not significant. **c**, Representative high dimensional spectral flow cytometry analysis in participant with severe obesity. Flow cytometric plots of RBD-binding (Top panels) and Spike-binding (Lower panels) B cells (CD19+) in a patient with severe obesity, prior to (V3D0), eight days after (V3D8) and 28 days after (V3D28) a booster mRNA vaccine. **d**, High-dimensional spectral flow cytometry of SARS-CoV-2 Spike-binding B cells. t-distributed stochastic neighbor embedding (tSNE) and FlowSOM (Flow Self Organizing Maps tool) analyses of multi-parameter flow cytometry of CD19+ Spike-binding B cells from normal weight individuals (Top panels) and participants with severe obesity (Lower panels) prior to (V3D0), eight days after (V3D8) and 28 days after (V3D28) a booster mRNA vaccine. Heatmap of cell surface expression of proteins on different clusters of cells; red indicates high expression, and yellow indicates low expression. **e**, Flow cytometric quantification of Spike-binding B cells that have been activated by the vaccine (IgD-) in normal weight individuals (blue) and participants with severe obesity (green) eight days (V3D8) and 28 days after (V3D28) a booster mRNA vaccine. Each symbol represents an individual person, horizontal bar indicates the median and p-values are from a Mann-Whitney U test.

**Supplementary Figure 4:**
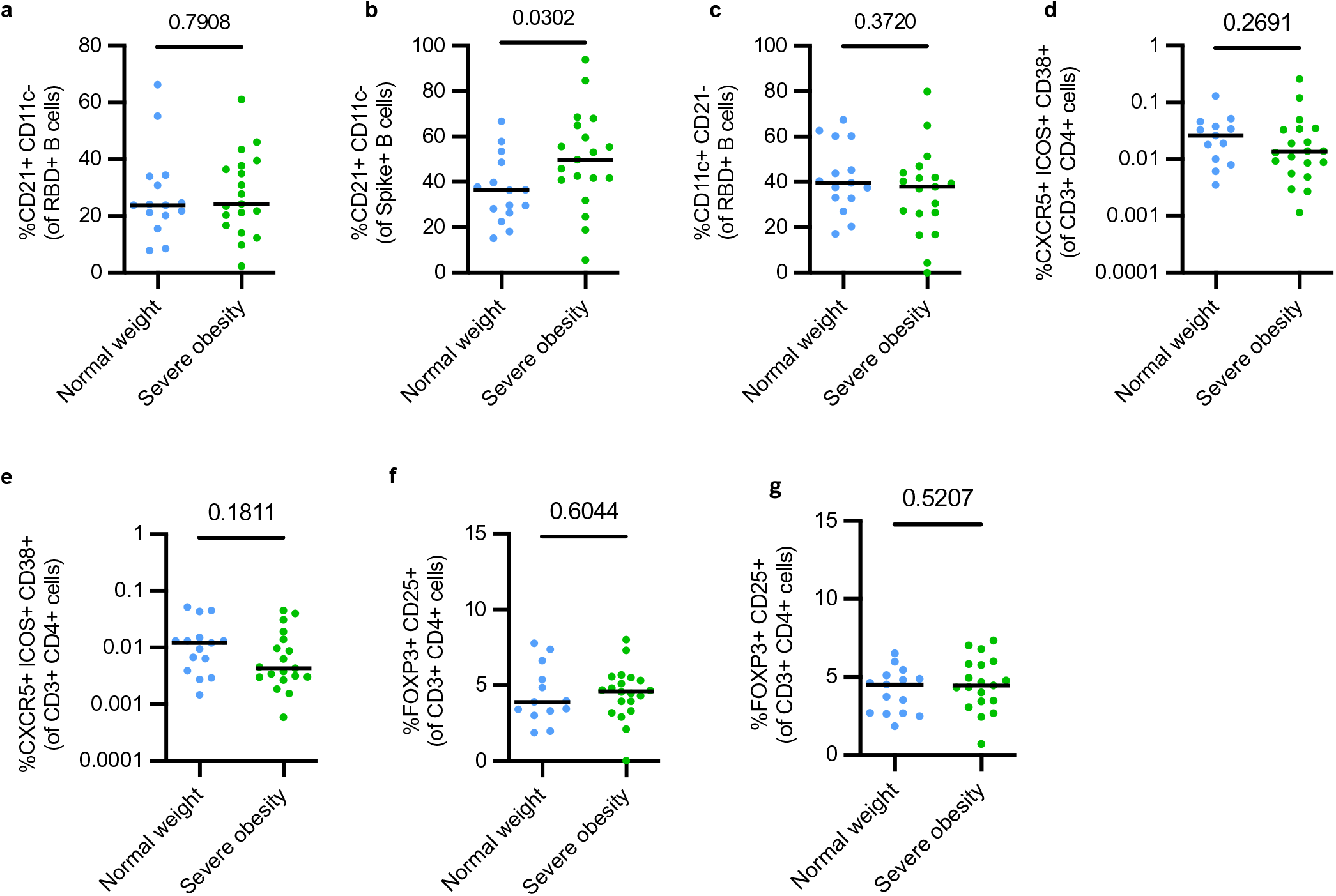
T and B cell response to third dose booster vaccination in severe obesity **a-c**, Flow cytometric quantification of RBD-binding or Spike-binding B cells of either a classical memory (**a** & **b**) or atypical memory (**c**) phenotype in normal weight individuals (blue) and participants with severe obesity (green) 28 days after (V3D28) a third dose mRNA vaccine. Each symbol represents an individual person, horizontal bars indicate the median and p-value is from a Mann-Whitney U test. **d-e**, Flow cytometric quantification of circulating T follicular helper (cTFH) cells (CXCR5+ ICOS+ CD38+ FOXP3- CD25- CD4+ cells) in normal weight individuals (blue) and participants with severe obesity (green) 8 days (**d**) and 28 days (**e**) after a third dose mRNA vaccine. Each symbol represents an individual person, horizontal bars indicate the median and p-value is from a Mann-Whitney U test. **f-g**, Flow cytometric quantification of circulating T regulatory cells (FOXP3+ CD25+ CD4+ cells) in normal weight individuals (blue) and participants with severe obesity (green) 8 days (**f**) and 28 days (**g**) after a third dose mRNA vaccine. Each symbol represents an individual person, horizontal bars indicate the median and p-value is from a Mann-Whitney U test.

**Supplementary Figure 5:**
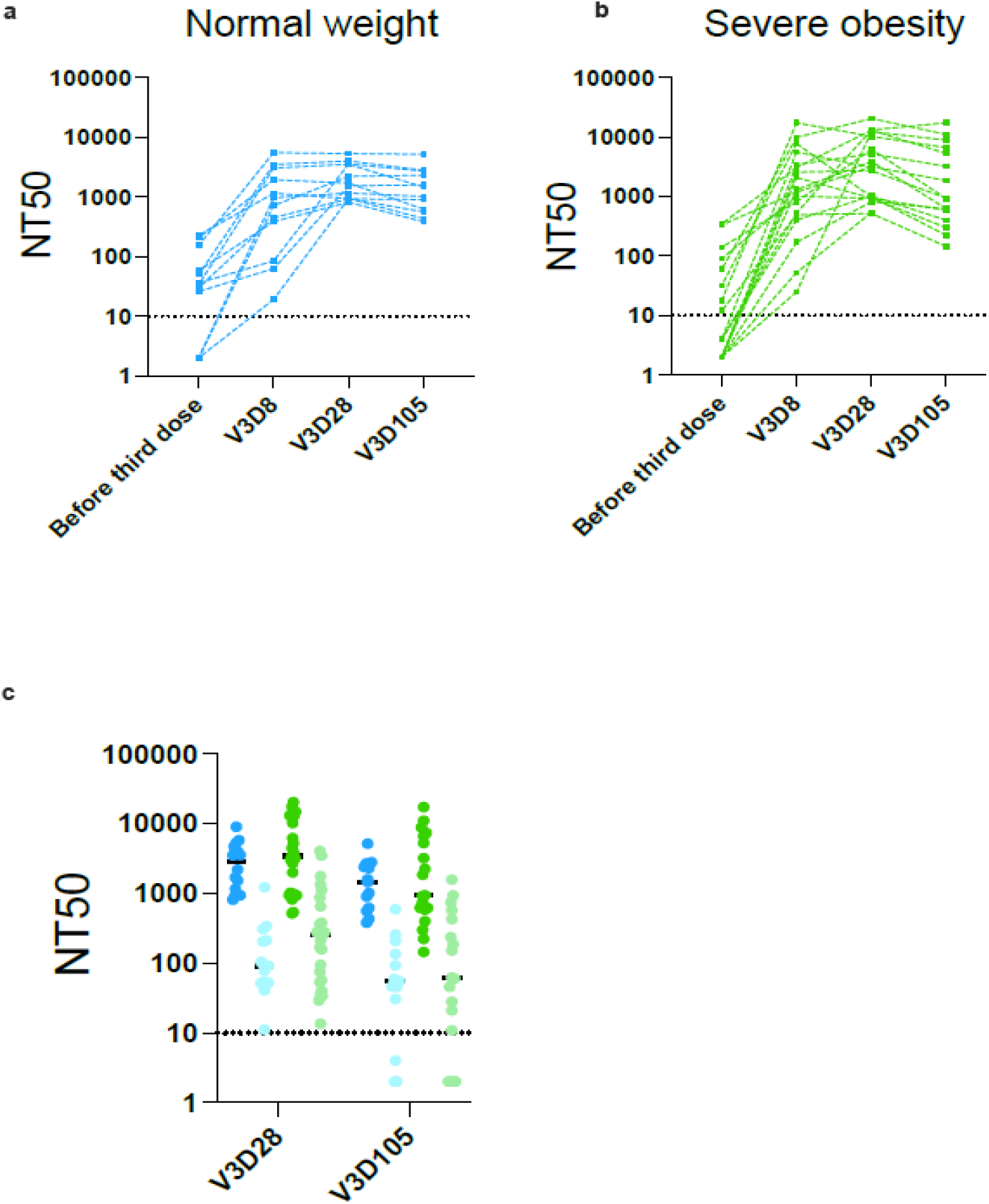
Neutralizing capacity before and after third vaccine dose. **a, b**, Neutralizing capacity (NT50) measured before and 8 days (V3D8), 28 days (V3D28) and 105 days (V3D105) after third dose vaccination (V3) in normal weight controls shown in blue **(a)** and people with severe obesity shown in green (**b)**. People who reported a positive SARS-CoV-2 test or had positive N antibodies at any time point were excluded. **c**, NT50 against wild-type SARS-CoV-2 (dark blue and dark green symbols) and Omicron (BA.1) variant (light blue and light green symbols) at day 28 and day 105 post third dose. Each symbol represents an individual person, horizontal bars indicate the median.

**Supplementary Figure 6:**
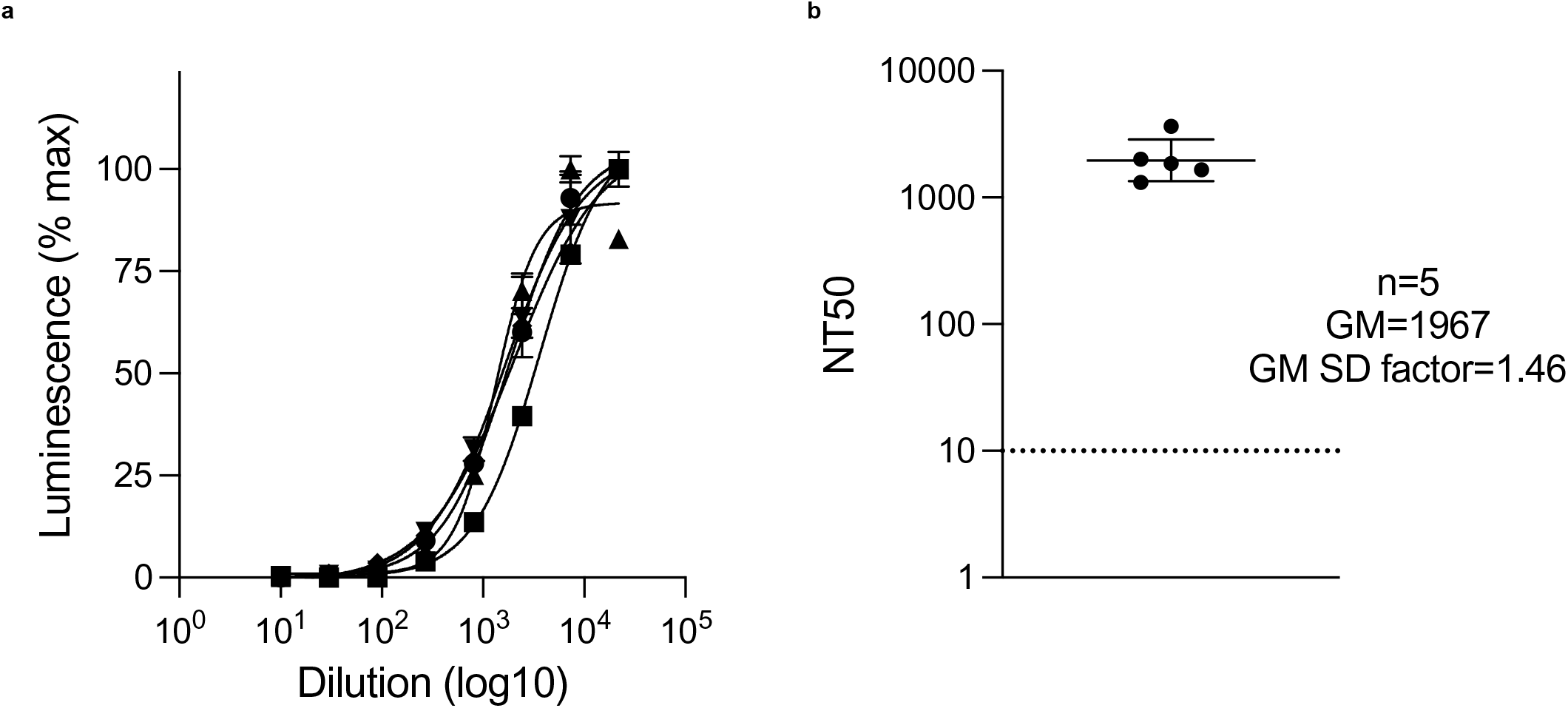
Neutralizing capacity of WHO International Standard 20/136. Neutralization curves **(a)** and corresponding NT50s **(b)** for WHO International Standard for anti-SARS CoV-2 immunoglobulin (IS 20/136) against wildtype SARS-CoV-2. Data from 5 independent experiments. Viral stock (MOI=0.01) was pre-incubated with a 3-fold dilution series of heat-inactivated standard before addition to luminescent reporter cells. Firefly luciferase activity was measured by luminometry 16 h post-infection. For each dilution, mean luminescence ± Standard Error of the Mean is displayed as % maximum. Geometric mean (GM) NT50 ± 95% confidence intervals are shown, with the dotted line indicating the limit of quantitation.

**Supplementary Table 1:**
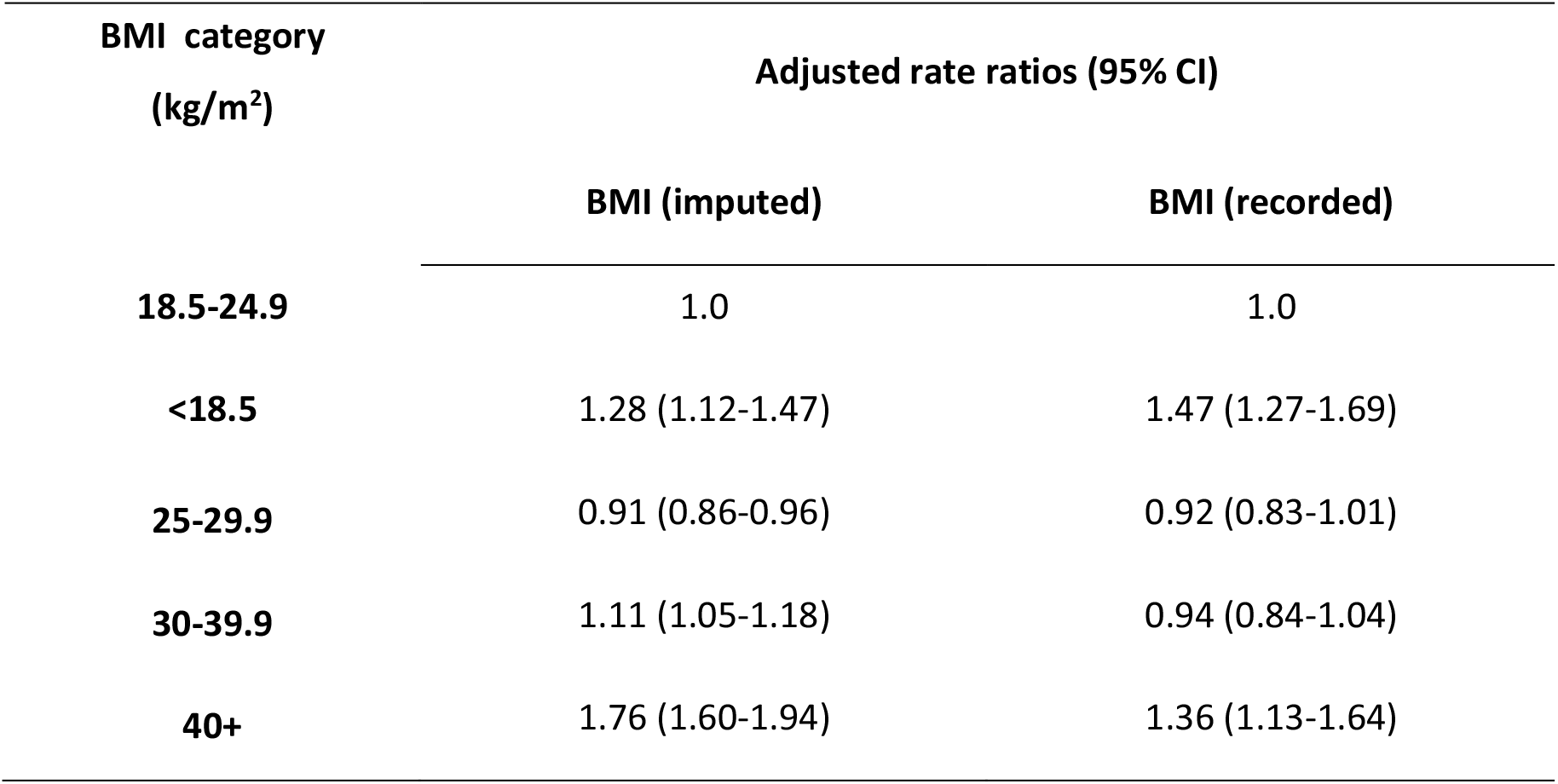
Association between BMI and Covid-19 hospitalization or death among individuals from the EAVE II cohort The frequency and rate per 1,000 person-years of severe Covid-19 outcomes (Covid-19 related hospitalization or death) was calculated. Adjusted rate ratios (aRRs) were estimated adjusting for all confounders including age, sex, Scottish Index of Multiple Deprivation, time since receiving the second dose of vaccine, number of pre-existing comorbidities, the gap between vaccine doses, previous history of SARS-CoV-2 infection and calendar time. Where the BMI was missing, it was imputed using ordinary least squares regression with all other independent variables included as predictors (BMI (imputed)). CI, confidence intervals. BMI=body mass index.

| BMI category<br>(kg/m<sup>2</sup>) | Adjusted rate ratios (95% CI) |  |
| --- | --- | --- |
|  | BMI (imputed) | BMI (recorded) |
| **18.5-24.9** | 1.0 | 1.0 |
| **<18.5** | 1.28 (1.12-1.47) | 1.47 (1.27-1.69) |
| **25-29.9** | 0.91 (0.86-0.96) | 0.92 (0.83-1.01) |
| **30-39.9** | 1.11 (1.05-1.18) | 0.94 (0.84-1.04) |
| **40+** | 1.76 (1.60-1.94) | 1.36 (1.13-1.64) |

**Supplementary Table 2:**
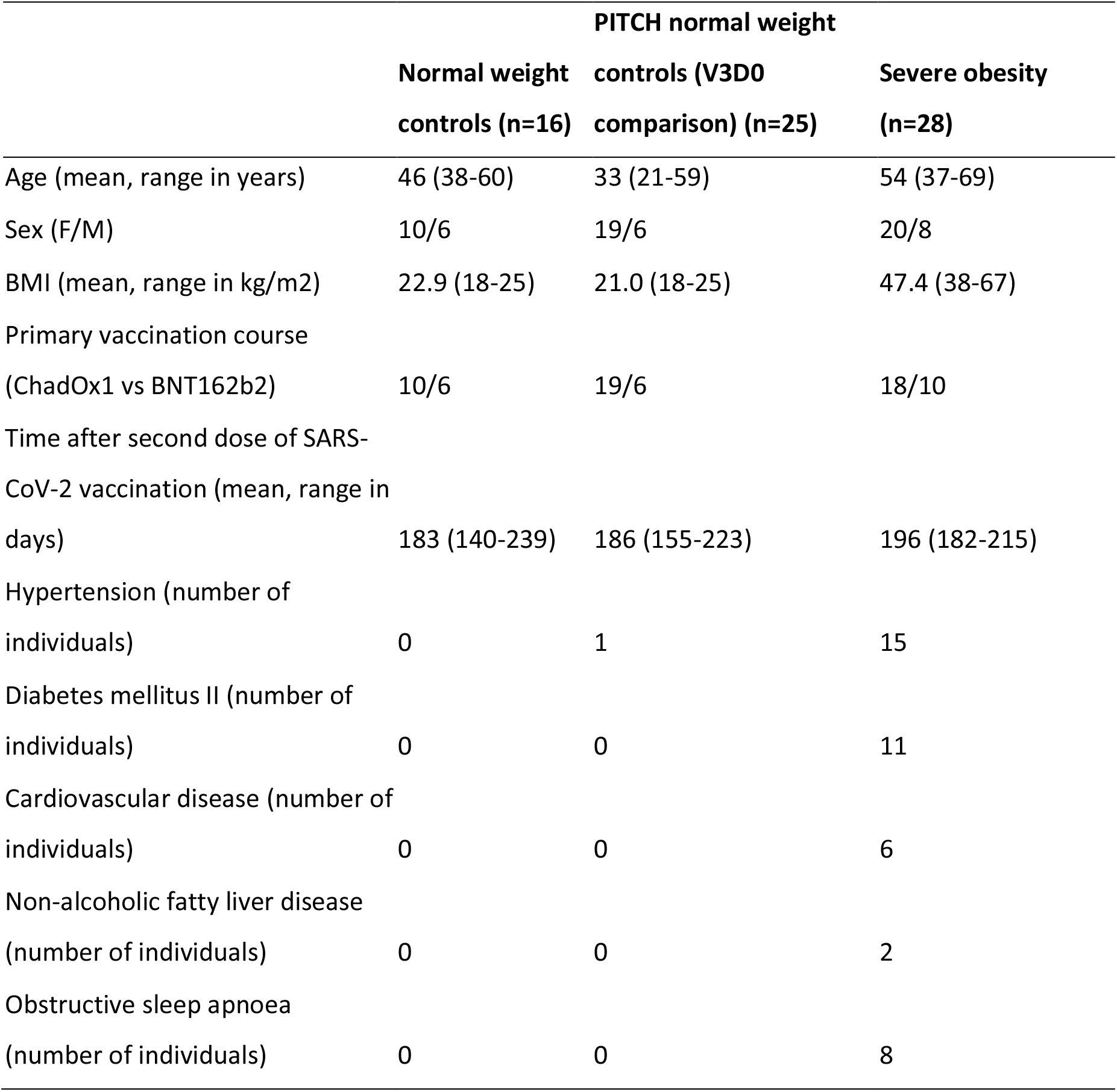
Characteristics of people with severe obesity and normal weight controls (SCORPIO study)

## Notes

### Competing Interest Statement

AS is a member of the Scottish Governments Standing Committee on Pandemic Preparedness and the Risk Stratification Subgroup of the UK Governments New and Emerging Respiratory Virus Threats Advisory Group (NERVTAG). He was a member of AstraZenecas Thrombotic Thrombocytopenic Task Force. All roles are unremunerated. SJD is a Scientific Advisor to the Scottish Parliament on COVID-19 for which she receives a fee. All other authors have no conflict of interest to declare.

### Funding Statement

The epidemiological study is part of the EAVE II project. EAVE II is funded by the MRC with the support of BREATHE The Health Data Research Hub for Respiratory Health which is funded through the UK Research and Innovation Industrial Strategy Challenge Fund and delivered through the Health Data Research UK. This research is part of the Data and Connectivity National Core Study led by Health Data Research UK in partnership with the Office for National Statistics and funded by UK Research and Innovation and the National Core Studies Immunity. Additional support has been provided through Public Health Scotland the Scottish Government Director-General Health and Social Care and the University of Edinburgh. The original EAVE project was funded by the National Institute for Health Research (NIHR) Health Technology Assessment programme.
The SCORPIO study was supported by the Medical Research Council (MR/W020564/1, a core award to J.E.T.; MC_UU_0025/12 and MR/T032413/1, an award to N.J.M.) and the Medical Research Foundation (MRF-057-0002-RG-THAV-C0798). Additional support was provided by NHSBT (WPA15-02 to N.J.M.), Addenbrookes Charitable Trust (900239 to N.J.M.) and the National Institute for Health Research (NIHR) Cambridge Biomedical Research Centre (N.J.M. and I.S.F.) and NIHR BioResource. M.A.L is supported by the Biotechnology and Biological Sciences Research Council (BBS/E/B/000C0427, BBS/E/B/000C0428) and is a Lister Institute Fellow and an EMBO Young Investigator. I.M.H. is supported by a CIMR PhD studentship; H.J.S. by a Sir Henry Dale Fellowship jointly funded by Wellcome and the Royal Society [109407] and a BBSRC institutional programme grant [BBS/E/B/000C0433]. I.S.F. is supported by Wellcome (207462/Z/17/Z), Botnar Fondation, the Bernard Wolfe Health Neuroscience Endowment and a NIHR Senior Investigator Award. The PITCH study was funded by the UK Department of Health and Social Care. S.J.D. is funded by an NIHR Global Research Professorship (NIHR300791). P.K. is an NIHR Senior Investigator and is funded by Wellcome (WT109965MA).

### Author Declarations

Ethical approval was granted by the National Research Ethics Service Committee, Southeast Scotland 02 (reference number: 12/SS/0201) for the study using the Early Pandemic Evaluation and Enhanced Surveillance of Covid-19 (EAVE II) platform. Clinical studies in people with severe obesity and normal weight controls were approved by the National Research Ethics Committee and Health Research Authority (East of England, Cambridge Research Ethics Committee (SCORPIO study, SARS-CoV-2 vaccination response in obesity amendment of NIHR BioResource 17/EE/0025)). Additional normal weight controls were recruited in Oxford, UK as part of the PITCH study under the GI Biobank Study 16/YH/0247, approved by Yorkshire & Humber Sheffield Research Ethics Committee.

